# Unveiling Peripheral Immune Dysfunction in Parkinson’s Disease through Analysis of Blood-based Mitochondrial DNA Copy Number

**DOI:** 10.1101/2024.04.28.24306517

**Authors:** Longfei Wang, Jiru Han, Liam G. Fearnley, Michael Milton, Haloom Rafehi, Joshua Reid, Zachary Gerring, Shashank Masaldan, Tali Lang, Terence P. Speed, Melanie Bahlo

## Abstract

Mitochondrial dysfunction plays an important role in Parkinson’s disease (PD), with mitochondrial DNA copy number (mtDNA-CN) emerging as a potential marker for mitochondrial health. Our study aimed to assess the association between blood mtDNA-CN and PD, as well as to uncover the underlying mechanisms. Introducing mitoCN, a novel mtDNA-CN estimator adjusting for coverage bias, suitable for large-scale whole-genome sequencing data, we applied it across six cohorts within the Accelerating Medicines Partnership program for Parkinson’s Disease dataset. We investigated the links between blood mtDNA-CN and both PD risk and severity, leveraging comprehensive clinical assessments. Our findings reveal that reduced blood mtDNA-CN levels are associated with heightened PD risk and increased severity of motor symptoms and olfactory dysfunction. However, upon adjusting for blood composition, these associations largely disappeared, indicating a predominant influence of changes in blood variables. Furthermore, using bidirectional Mendelian randomization, we explored causal relationships, finding no evidence of a direct causal relationship between blood mtDNA-CN and PD susceptibility. Thus, even though blood bulk mtDNA-CN correlates with an elevated risk of PD and more severe PD symptoms, our refined analyses and results suggest that peripheral immune dysfunction rather than mitochondrial dysfunction underpins these previously identified associations.

## Introduction

Parkinson’s disease (PD) is the second most common neurodegenerative disorder worldwide. According to the World Health Organization (WHO), 8.5 million individuals live with PD globally in 2019. PD is reported to result in a loss of 5.8 million disability-adjusted life years (DALYs), with a reported increase of 81% in morbidity since 2000. PD is characterized by the progressive degeneration of dopaminergic neurons in the substantia nigra region of the brain, resulting in a deficiency of the neurotransmitter dopamine and causing motor symptoms like tremors, bradykinesia, and rigidity, as well as contributing to non-motor symptoms such as sleep disturbances, anosmia, and cognitive impairment^1^. Notably, mitochondrial dysfunction is recognized as a significant contributor to the pathogenesis of PD^2^. Mitochondria serve as the cellular powerhouses responsible for generating adenosine 5′ triphosphate (ATP) by oxidative phosphorylation. In PD, dysfunctional mitochondria can lead to impaired energy production, oxidative stress, and molecular damage, which are believed to contribute to the death of dopaminergic neurons and disease progression^3^.

The diagnosis of PD primarily relies on clinical assessment of motor symptoms, which can result in delays in diagnosis or even misdiagnosis. Early detection of PD is crucial because current pharmacological treatments are more effective when initiated in the early stages of the disease. Additionally, lifestyle changes such as increased physical activity have greater efficacy during the early stages of the disease, as individuals typically experience milder motor symptoms and can engage in more physical exercises without risking falls^4^. Therefore, there is a need for reliable and non-invasive biomarkers capable of assessing mitochondrial health, thus facilitating the early diagnosis of PD.

Mitochondrial DNA copy number (mtDNA-CN), referring to the quantity of mitochondrial DNA (mtDNA) molecules present in cells, is a readily measurable indicator of mitochondrial function. mtDNA-CN can be measured using laboratory techniques like quantitative real-time polymerase chain reaction (qPCR) as well as high throughput methods such as genotyping arrays, whole genome sequencing (WGS), and whole exome sequencing (WES)^5^. Employing the qPCR technique on a cohort of 363 peripheral blood samples and 151 substantia nigra pars compacta tissue samples, Pyle and colleagues observed a significant reduction in mtDNA-CN in both peripheral blood and substantia nigra of individuals with PD when compared to matched controls^6^. Several studies which estimated mtDNA-CN using sequencing data from the UK Biobank (UKB)^7^ have discovered statistically significant associations between blood mtDNA-CN and common diseases, including PD^8–10^. Despite multiple reported links between mtDNA-CN and PD, to our knowledge, no study has investigated this relationship in large-scale, PD-specific cohorts with deeper, PD-specific clinical assessments, which the UKB lacks.

Longchamps et al. evaluated the existing mtDNA-CN estimation techniques and determined that mtDNA-CN derived from WGS is the most reliable choice for capturing association signals^5^. Existing WGS-derived mtDNA-CN estimators calculate the ratio of the mean coverage of mtDNA to the mean coverage of nuclear DNA (nucDNA), assuming that short reads are uniformly distributed in their alignment throughout the whole genome^8,9,11^. However, coverage bias, which stems from factors like GC content and homology, can introduce unwanted variation in the mean coverage of both mtDNA and nucDNA^12,13^. With the rapid increase in the availability of WGS data, more accurate and efficient methods are required.

In this study, we introduce a novel mtDNA-CN estimator, named mitoCN, designed to quantify mtDNA-CN from WGS data while accounting for GC bias and homology adjustments. Using data from the cohorts available through the Accelerating Medicine Partnership program for Parkinson’s Disease (AMP PD) portal, we investigated the association between blood mtDNA-CN and PD risk, as well as motor and non-motor clinical assessments. We found that lower blood mtDNA-CN is associated with a higher PD risk and more severe motor symptoms and olfactory impairment. Additionally, we revealed that these associations are predominantly due to changes in blood markers of immune system function. To validate our findings, we replicated our analysis using the WGS data of approximately 500,000 participants from the UKB.

## Results

### mtDNA-CN estimation

In the discovery study, we utilized the AMP PD v3 dataset, released on November 15, 2022. AMP PD consolidates data from eight unified cohorts: BioFIND, Harvard Biomarkers Study (HBS), Lewy body dementia case-control cohort (LBD), LRRK2 Cohort Consortium (LCC), Parkinson’s disease Biomarkers Program (PDBP), Parkinson’s Progression Markers Initiative (PPMI), Study of Isradipine as a Disease-modifying Agent in Subjects With Early Parkinson Disease, Phase 3 (STEADY-PD3), and the Study of Urate Elevation in Parkinson’s Disease, Phase 3 (SURE-PD3). AMP PD provides comprehensive clinical data for all participants, along with WGS data for 10,418 jointly genotyped samples as well as transcriptomics data for 3,274 participants, including 8,461 whole blood bulk RNA samples. Given our focus on PD in this study and the unavailability of sequence alignment data for the LCC cohort, we excluded the LBD and LCC cohorts from the analysis. AMP PD is housed on Terra, a cloud computing platform.

We developed mitoCN (https://github.com/bahlolab/mitoCN), a novel method designed to estimate mtDNA-CN from WGS data that adjusts for GC bias and homology bias (**Methods**). We assessed the concordance and percentage change between mtDNA-CN estimates produced by mitoCN and a recently published method called mtSwirl^9^ (**Methods**), which calculates mtDNA-CN using the ratio of the mean coverage of mtDNA to the mean coverage of nucDNA. Our analysis revealed high concordance in mtDNA-CN estimates between mitoCN and mtSwirl (R² = 0.999, *p* <0.0001, Supplementary Fig. 1A). Compared to mitoCN, the average percent change with mtSwirl for whole blood samples was -0.4%, closely approximating zero (Supplementary Fig. 1B). The range of the percent change extends from -21.7% to 15.2%. mtSwirl demonstrated slightly improved mtDNA coverage among African and East Asian groups by constructing self-reference sequences for each sample (Supplementary Fig. 1C). The percent change in mtDNA-CN was 0.84% (range: -2.79% to 3.62%) for Africans and 0.03% (range: -3.59% to 2.84%) for East Asians. While mtSwirl is designed for calling both mtDNA variants and copy number by constructing self-reference sequences, mitoCN focuses solely on mtDNA-CN estimation using aligned reads, offering faster computational speeds compared to mtSwirl. For example, it requires around 10 minutes of CPU time for a 30X genome. Given the emphasis of our study on mtDNA-CN estimation, we opted for mitoCN for our analysis.

Even though DNA source information for WGS samples was consistently recorded as “whole blood” in AMP PD, we identified two distinct distributions in the mtDNA-CN estimates (Supplementary Fig. 2A). This observation suggests that DNA samples were extracted from two types of blood samples. Given these independent distributions, it is necessary to analyze the data from these two DNA sources separately. Therefore, we classified the samples into two clusters using a Gaussian mixture model^14^ (Supplementary Fig. 2B). Platelets play a significant role in the discrepancy of the mtDNA-CN distributions, as they exclusively contain mtDNA without nucDNA^15^. A previous study showed that the estimation of mtDNA-CN from whole blood samples is twice as high as that from leukocytes^16^. Consequently, we interpreted cluster 1 as comprising platelet-depleted blood samples, such as leukocytes or peripheral blood mononuclear cells (PBMCs), and cluster 2 as containing platelet-abundant blood samples, such as those derived from whole blood or buffy coat samples. Cohorts differed markedly in their contributions to these two clusters with the PDBP cohort having 84.2% of samples from cluster 1 (also referred to as “platelet-depleted blood samples”) with the remaining samples allocated to cluster 2 (also referred to as “platelet-abundant blood samples”). In contrast, cohort PPMI had 39.7% samples from cluster 1 (Table 1).

**Table 1.** Description of the AMP PD datasets and mtDNA-CN clustering results.

| Study | Samples | Female (%) | Diagnosis at baseline |  |  | Known PD mutation carriers | WB bulk RNA-seq | Age: mean (SD) |
| --- | --- | --- | --- | --- | --- | --- | --- | --- |
|  |  |  | Case | Control | Other |  |  |  |
| Cluster 1: platelet-depleted blood samples |  |  |  |  |  |  |  |  |
| BioFIND | 172 | 71 (41%) | 99 | 70 | 3 | 48 (28%) | 167 | 67 (6.9) |
| PDBP | 1,263 | 558 (44%) | 728 | 432 | 103 | 389 (31%) | 1,232 | 64 (10.1) |
| PPMI | 718 | 258 (36%) | 465 | 224 | 29 | 286 (40%) | 647 | 62 (10.7) |
| Cluster 2: platelet-abundant blood samples |  |  |  |  |  |  |  |  |
| HBS | 1,173 | 580 (49%) | 639 | 531 | 3 | 350 (30%) | 0 | 67 (10.2) |
| PDBP | 237 | 104 (44%) | 130 | 69 | 38 | 72 (30%) | 148 | 65 (9.7) |
| PPMI | 1,089 | 576 (53%) | 467 | 540 | 82 | 946 (87%) | 799 | 61 (11.7) |
| STEADY-PD3 | 329 | 100 (30%) | 329 | 0 | 0 | 101 (31%) | 0 | 62 (9.2) |
| SURE-PD | 259 | 127 (49%) | 259 | 0 | 0 | 90 (35%) | 0 | 63 (9.6) |
| **Total** | 5,240 | 2,374 (45%) | 3,116 | 1,866 | 258 | 2,282 (44%) | 2,993 | 64 (10.6) |

WB = whole blood; SD = standard deviation; Other = other diagnosis, including prodromal PD, Alzheimer's disease, Lewy body dementia, and other.

### Investigating associations between blood mtDNA-CN and PD

Previous studies have consistently reported that blood mtDNA-CN tends to be lower in males compared to females and declines with age^5,17^. We were able to confirm these previous findings with mtDNA-CN through individual cohort analyses and meta-analyses (**Methods**, Supplementary Figs. 3 - 4).

To investigate the association between blood mtDNA-CN and the risk of PD, we first performed cohort analysis by fitting robust linear models using the “rlm()” function in the MASS R package, with PD diagnosis at baseline, while adjusting for age, sex, and ancestry using the first five principal components (PCs) (**Methods**, Supplementary Table 1A). Significant associations were observed in the PDBP cohort (*p* = 0.008) within cluster 1, and in the HBS (*p* = 0.01) and PPMI (*p*

<0.0001) cohorts within cluster 2, indicating that individuals diagnosed with PD have lower mtDNA-CN compared to healthy controls. The STEADY-PD3 and SURE-PD cohorts only included PD cases and were therefore not included in this analysis. Subsequently, using the summary statistics from the cohort analysis, we performed a meta-analysis for each cluster (**Methods**, Fig. 1). We observed a significant association in cluster 1 (beta: -0.03, 95% confidence interval [CI]: [-0.05, -0.01], *p* = 0.006), but not in cluster 2, owing to the heterogeneity among the cohorts (Cochran’s Q = 7.99, *p* = 0.02). Upon excluding the PDBP cohort from cluster 2, there was no evidence of heterogeneity between the PPMI and HBS cohorts (Cochran’s Q = 0.41, *p* = 0.52), and the association becomes significant (beta = -0.10, 95% CI: [-0.13, -0.07], *p* <0.0001). Overall, our findings demonstrate that lower blood mtDNA-CN is associated with a higher risk of PD, regardless of the likely origin of the cell type of the samples (“platelet abundant” or “platelet non-abundant”).

**Fig. 1.**
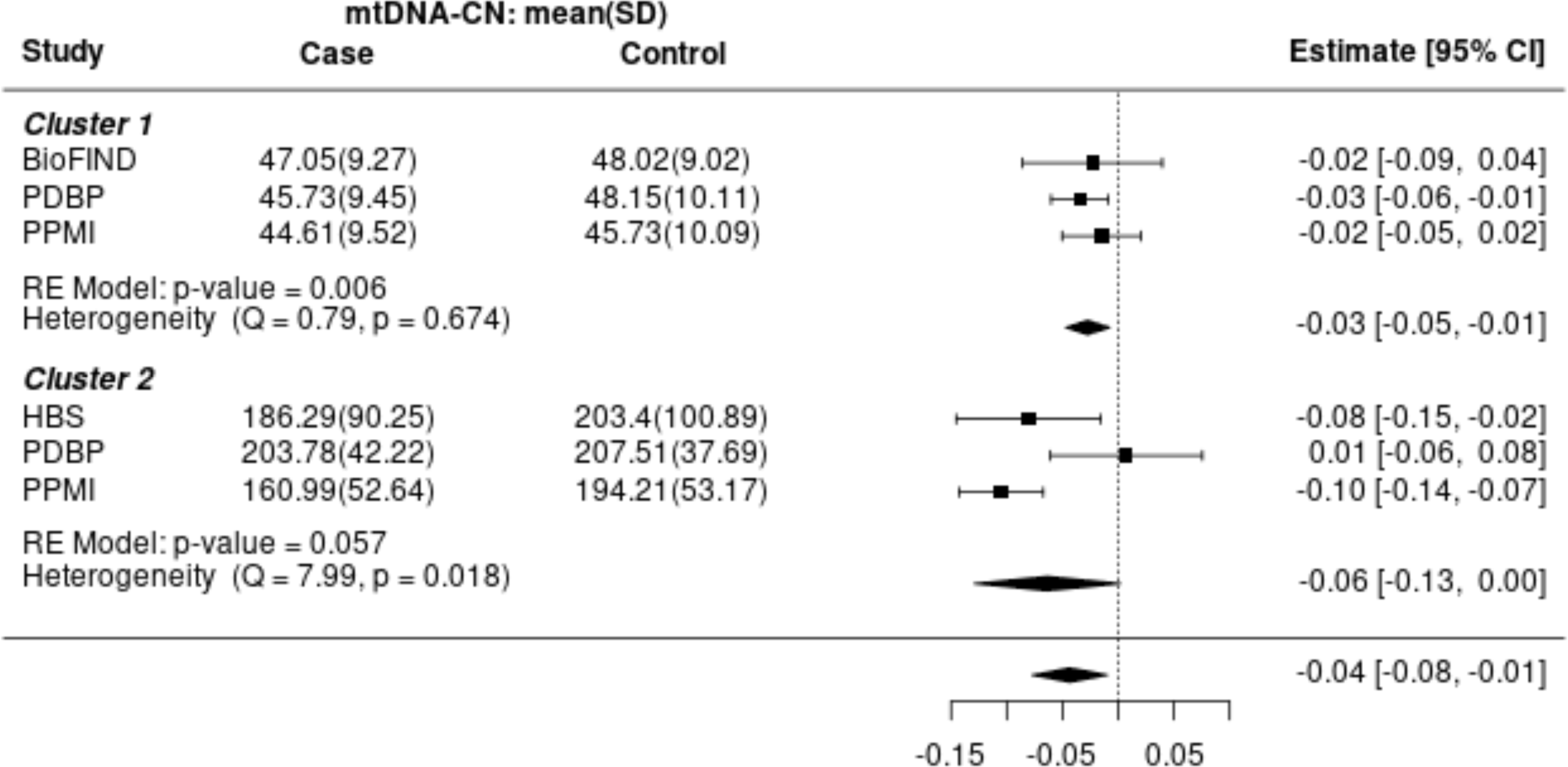
Meta-analysis of association tests between mtDNA-CN and PD diagnosis. For cluster 1, the meta-analysis results indicate that individuals diagnosed with PD have lower mtDNA-CN compared to healthy controls (beta = -0.03, *p* = 0.006). However, the association is not significant in cluster 2, mainly due to the heterogeneity induced by the PDBP cohort. Upon excluding the PDBP cohort, there is no evidence of heterogeneity between the PPMI and HBS cohorts (Q = 0.41, *p* = 0.52), and the association becomes significant (beta = -0.10, *p* <0.0001).

We explored the associations of mtDNA-CN with the severity of PD using several clinical assessments provided by AMP PD, including the Movement Disorder Society-sponsored revision of the Unified Parkinson’s Disease Rating Scale part III (MDS-UPDRS III) for clinical motor examination, activities of daily living (ADL), olfactory impairment as assessed by the University of Pennsylvania Smell Identification Test (UPSIT), and cognitive performance measured by the Montreal Cognitive Assessment (MoCA). Our analysis revealed significant associations of mtDNA-CN with MDS-UPDRS III (cluster 1: beta = -0.003, p <0.0001; cluster 2: beta = -0.003, p <0.0001), ADL (cluster 1: beta = 0.002, p = 0.02; cluster 2: beta = 0.002, p <0.0001), and UPSIT (cluster 1: beta = 0.002, p <0.0001; cluster 2: beta = 0.005, p <0.0001) scores across both clusters (Supplementary Figs. 5 - 7). However, we observed no significant association between mtDNA-CN and MoCA score, except within the PPMI cohort (Supplementary Fig. 8). Notably, the olfactory bulb has been identified as one of the first regions of insult in PD^18^, and the UPSIT score has been linked to the severity of PD. In summary, these findings indicate that decreased blood mtDNA-CN levels are associated with increased severity of motor symptoms and olfactory dysfunction, but not with cognitive decline.

Blood bulk mtDNA-CN is known to be influenced by blood composition, as reported in previous studies^19,20^. Gupta et al.^9^ suggested that previously reported associations between low blood mtDNA-CN and increased risk of common diseases are secondary to changes in blood composition. Therefore, it is important to determine whether the observed associations could also be attributed to changes in cell composition. To address this, we require a complete cell count profile from the blood samples used for DNA extraction. Unfortunately, blood cell measurements are not available in the AMP PD dataset. To overcome this limitation, we estimated cell type proportions from the whole blood bulk RNA-sequencing (RNA-seq) data using CIBERSORTx^21,22^ and the LM22 signature matrix^23^, a well-established reference for PBMCs (**Methods**). For this analysis, we utilized a subset of the PPMI cohort, which includes the most comprehensive clinical data and whole blood bulk RNA-seq data collected at baseline (N = 785). Using stepwise regression model selection, we included six blood cell proportions for blood composition correction, namely naïve B cells, naïve CD4 T cells, resting memory CD4 T cells, resting mast cells, neutrophils, and white blood cells. Importantly, all the selected blood variables are markers of immune system function. The adjusted mtDNA-CN measure was defined as the residual of the log-scaled raw mtDNA-CN with cell composition correction (**Methods**).

Subsequently, we examined the associations of PD-related variables with both raw and adjusted mtDNA-CN (**Methods**, Table 2). Lower raw mtDNA-CN is associated with a higher risk of PD, increased severity in motor experiences of daily living (MDS-UPDRS II), motor examination (MDS-UPDRS III), cognitive decline (MoCA), and olfactory impairment (UPSIT). Correction for blood cell composition attenuated the mtDNA-CN PD effect size estimates; however, the associations for PD risk and olfactory impairment remained significant after multiple testing correction. Notably, the UPSIT score demonstrates the most significant association with both raw and adjusted mtDNA-CN (t = 4.76, p <0.0001, and t = 3.56, p = 0.005), respectively. In conclusion, changes in blood composition, reflecting the peripheral immune dysfunction in individuals with PD, partially explain the association between mtDNA-CN and PD. The remaining signal may arise from the limitation of the CIBERSORTx estimates.

**Table 2.** Association test results of raw and adjusted mtDNA-CN with PD variables.

| PD variable | sample size | raw mtDNA-CN |  |  |  | adjusted mtDNA-CN |  |  |  |
| --- | --- | --- | --- | --- | --- | --- | --- | --- | --- |
|  |  | beta | t | p | p.adj | beta | t | p | p.adj |
| diagnosis | 726 | -0.108 | -4.54 | <0.0001 | <0.0001 | -0.072 | -3.05 | 0.002 | 0.02 |
| MDS UPDRS I | 785 | 0.002 | 1.02 | 0.31 | 0.49 | 0.003 | 1.48 | 0.14 | 0.33 |
| MDS UPDRS II | 785 | -0.007 | -3.80 | 0.0002 | 0.0006 | -0.004 | -2.25 | 0.03 | 0.09 |
| MDS UPDRS III | 785 | -0.004 | -4.65 | <0.0001 | <0.0001 | -0.002 | -2.55 | 0.01 | 0.05 |
| MDS UPDRS IV | 214 | 0.011 | 1.64 | 0.10 | 0.20 | 0.007 | 1.11 | 0.27 | 0.41 |
| MoCA | 784 | 0.011 | 3.13 | 0.002 | 0.005 | 0.006 | 1.72 | 0.09 | 0.24 |
| RBD | 781 | 0.002 | 0.60 | 0.55 | 0.59 | 0.000 | -0.09 | 0.93 | 0.93 |
| ESS | 514 | -0.003 | -1.00 | 0.32 | 0.49 | -0.004 | -1.21 | 0.23 | 0.40 |
| UPSIT | 768 | 0.006 | 4.76 | <0.0001 | <0.0001 | 0.005 | 3.56 | 0.0004 | 0.005 |
| ADL | 756 | 0.002 | 2.17 | 0.03 | 0.07 | 0.001 | 0.60 | 0.55 | 0.70 |
| MRI | 364 | 0.022 | 0.79 | 0.43 | 0.56 | 0.033 | 1.20 | 0.23 | 0.40 |

The table shows the PD-related variables, sample sizes, effect sizes, t values, p values, and false discovery rate (FDR) adjusted p values for the output of multivariable regression models, modeling raw and adjusted mtDNA-CN on PD variable + age + sex + population, respectively. MDS UPDRS = Movement Disorder Society-Sponsored Revision of the Unified Parkinson's Disease Rating Scale; MoCA = Montreal Cognitive Assessment; RBD = Rapid eye movement sleep behavior disorder; ESS = Epworth Sleepiness Scale; UPSIT = University of Pennsylvania Smell Identification Test; ADL = activities of daily living; MRI = magnetic resonance imaging.

We then examined the relationship between PD risk and blood markers indicative of immune system function, including lymphocyte percentage, neutrophil percentage, and neutrophil-to-lymphocyte ratio (NLR). Lymphocyte percentage was calculated by summing T cells, B cells, and natural killer (NK) cells. Multivariable robust linear models were used, modeling blood markers on PD variable + age + sex + population (PC1-5). All three blood markers were significantly associated with both risk and severity of PD, as assessed by MDS UPDRS I-III, ADL, and UPSIT scores (Supplementary Table 1B), even after false discovery rate (FDR) multiple testing correction.

Using the GWAS summary statistics for both raw and adjusted blood mtDNA-CN from Gupta et al., 2023^9^, we constructed the polygenic risk scores (PRSs) per individual in AMP PD (Supplementary Table 1C). Subsequently, we assessed the relationships between the actual mtDNA-CN estimates and these PRSs. Despite observing significantly positive correlations between blood mtDNA-CN estimates and PRSs in most cohorts, the correlation coefficients were found to be modest (Supplementary Table 1D and Supplementary Fig. 9). This modest correlation can be attributed to the fact that mtDNA-CN is influenced by both genetic and environmental factors, with a SNP-based heritability of approximately 4%^9^. Additionally, the PRSs exhibit no correlation with mtDNA-CN estimates from brain samples (Supplementary Table 1D). Furthermore, our investigation into the associations between mtDNA-CN PRSs and PD did not reveal any significant associations with PD (raw mtDNA-CN PRS: *p* = 0.37, adjusted mtDNA-CN PRS: *p* = 0.65, see Supplementary Fig. 10).

### Investigating causal relationships between blood mtDNA-CN and PD

We performed bidirectional two-sample Mendelian Randomization (MR) analyses using GWAS summary statistics to assess the causal relationship between the risk of PD and blood mtDNA-CN estimated from different sequencing data, WGS, genotyping arrays, and a combination of WES and genotyping array data (Supplementary Table 1C). The inverse variance-weighted (IVW) method analysis showed weak evidence for potential causal effects between mtDNA-CN and PD (Fig 2). The weighted median, weighted mode, and MR Egger regression approaches yielded similar estimates (Supplementary Table 1E). In summary, our findings suggest no direct causal relationship between blood mtDNA-CN and the risk of PD.

**Fig. 2:**
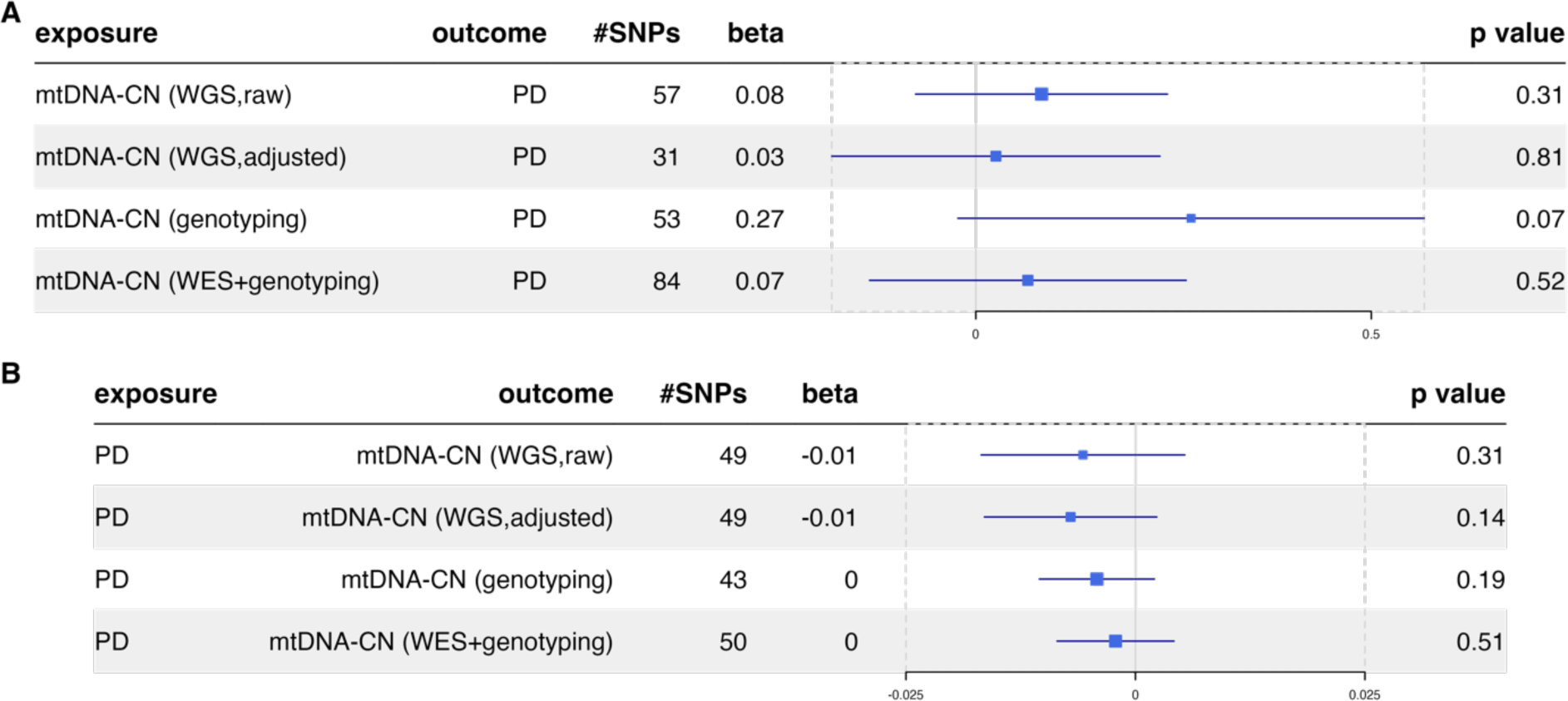
Mendelian randomization using the IVW method estimated the causal effects between blood-derived mtDNA-CN and PD susceptibility. **a** causal effect from mtDNA-CN estimates using different techniques to PD risk; **b** causal effects of PD on mtDNA-CN. The third column provides the number of SNPs selected as instrumental variables in the analysis. The forest plots visually represent the effect size, beta, along with the confidence interval.

The robustness of our findings was confirmed through sensitivity analysis (**Methods**). Cochran’s Q test was utilized to identify heterogeneity (Supplementary Table 1F). Given the detection of heterogeneity, we applied the random-effect IVW MR approach, ensuring the applicability of our results. Moreover, all intercepts assessed with the MR Egger method resulted in not significant p-values (Supplementary Table 1F), indicating that our results were not influenced by horizontal pleiotropy. In most cases, leave-one-out analyses did not identify SNPs that influenced the final estimates, and the funnel plots did not provide significant evidence of bias when evaluating potential biases in the genetic instrumental variables (IVs) (Supplementary Figs. 11 - 18).

### UK Biobank Replication Study

In the replication study, we utilized WGS data from approximately 500,000 participants in the UKB. This included data from the initial release of around 200,000 participants in late 2021, as well as data from the subsequent release of the remaining approximately 300,000 participants in late 2023. Participants were included only if they had complete data for all variables used in this analysis. We identified related individuals within third-degree relatedness (kinship coefficient >0.0625). To maximize the inclusion of PD cases, we excluded related individuals while ensuring the retention of as many cases as possible. In total, the analysis encompassed 367,322 samples (Table 3).

**Table 3.** Description of the UKB dataset.

| UKB | Sample size | Female (%) | PD cases | Mean age (SD) |
| --- | --- | --- | --- | --- |
| Batch 1 (2021 release) | 151,514 | 82,436 (54.4%) | 1,529 | 57 (8.1) |
| Batch 2 (2023 release) | 215,808 | 114,517 (53.1%) | 2,268 | 57 (8.1) |
| **Total** | 367,322 | 196,953 (53.6%) | 3,797 | 57 (8.1) |

The table displays sample sizes, female percentages, the number of PD cases, and the mean and standard deviation of age in the two batches of whole-genome sequencing data released by the UK Biobank.

The UKB provides a complete blood cell profile for the participants as measured with the Beckman Coulter LH750 instruments. We selected 21 blood measurements from the Category Blood Count (Category 100081), excluding nucleated red blood cell and reticulocyte measurements due to the absence of DNA in these cell types. Additionally, basophil measurements were excluded due to their low contributions to the total cell profile (<1%). Through stepwise regression model selection, the final statistical model incorporated nine blood measurements: white blood cell count, platelet count, plateletcrit (PCT), mean platelet volume, platelet distribution width, lymphocyte percentage, monocyte percentage, neutrophil percentage, and eosinophil percentage. Adjusted mtDNA-CN was determined using the residuals of the final model.

To assess the association of raw and adjusted blood mtDNA-CN with PD diagnosis, we applied robust linear models, modeling raw and adjusted mtDNA-CN on PD + sex + age + batch (2021 or 2023 release) + population (via first five ancestry PCs), respectively. Significant associations were observed in the raw mtDNA-CN (beta: -0.02, 95% CI: [-0.012, -0.028], *p <*0.0001, Table 4), indicating that individuals diagnosed with PD have lower mtDNA-CN compared to those without PD. However, following blood variable correction, no significant association was detected between mtDNA-CN and PD diagnosis (beta: -0.003, 95% CI: [0.004, -0.01], *p* = 0.38, Table 4), suggesting that the observed associations can largely be attributed to changes in blood composition. The associations between different blood variables and PD diagnosis are presented in Supplementary Table 1G. Following correction for multiple testing, we detected significant associations between PD risk and platelet count, PCT, as well as the count and percentage of lymphocytes, monocytes, neutrophils, and eosinophils, alongside the NLR. Notably, among these associations, NLR exhibits the largest effect with a t value of 12.37 and *p* <0.0001.

**Table 4.** Association test results of raw and adjusted blood mtDNA-CN with PD risk in the UKB cohort.

| Variables | raw mtDNA-CN |  |  |  | adjusted mtDNA-CN |  |  |  |
| --- | --- | --- | --- | --- | --- | --- | --- | --- |
|  | beta | se | t | p | beta | se | t | p |
| Sex | -0.043 | 0.001 | -53.89 | <0.0001 | 0.002 | 0.001 | 3.41 | 0.001 |
| Age | -0.003 | 0 | -55.34 | <0.0001 | -0.002 | 0 | -42.91 | <0.0001 |
| Batch | 0.018 | 0.001 | 22.39 | <0.0001 | 0.016 | 0.001 | 21.96 | <0.0001 |
| diagnosis | -0.020 | 0.004 | -5.07 | <0.0001 | -0.003 | 0.003 | -0.87 | 0.38 |

The table shows the effect sizes, standard errors, t values, and p values for the output of multivariable regression models, modeling raw and adjusted mtDNA-CN on PD + age + sex + batch + population, respectively. Baselines: sex - female; batch - batch1(2021 release); diagnosis - control.

## Discussion

While studies have previously utilized UKB data to investigate the association between mtDNA-CN and common disorders, including PD, none have utilized large-scale PD-specific data with comprehensive clinical assessments. By leveraging WGS data across multiple cohorts from the AMP PD resource, we report associations between blood bulk mtDNA-CN and both the risk and severity of PD. However, by correcting for both estimated and measured blood cell composition, we show that these associations can be predominantly attributed to blood markers of immune system function. Importantly, we demonstrate no causal relationship between blood mtDNA-CN and PD susceptibility using bidirectional two-sample MR.

While we did not find evidence of mitochondrial dysfunction through blood mtDNA-CN, our findings suggest that peripheral inflammatory immune responses may significantly contribute to the pathogenesis of PD. Previous studies have indicated that a lower lymphocyte count is associated with an increased risk of PD, driven by reductions in helper-CD4^+^ T cell and B-cell counts^24–27^. Additionally, several studies have suggested that higher neutrophil and lower lymphocyte counts are linked to an increased risk of PD and higher Unified Parkinson’s Disease Rating Scale (UPDRS) motor scores^28,29^. The neutrophil-to-lymphocyte ratio reflects the dynamic relationship between innate (neutrophils) and adaptive cellular immune response (lymphocytes) during illness and various pathological states^30^.

We recognized challenges in performing mtDNA-CN analysis, particularly regarding missing or incorrect DNA source information. The number of mitochondria varies across different tissues and cell types due to varying energy requirements and biological functions. For instance, tissues with higher energy demands such as the brain and liver exhibit higher mtDNA-CN. Additionally, platelets contain exclusively mtDNA but no nucDNA, resulting in higher mtDNA-CN levels in whole blood samples compared to PBMCs. Consequently, mtDNA-CN serves as a useful tool for identifying DNA cell type sources. When conducting mtDNA-CN analysis, it is crucial to ensure that the sequencing data are generated from the same DNA source or, at the very least account for this in the statistical modeling. The gnomAD resource has also identified this issue and chose to only include samples with mtDNA-CN falling within an arbitrary range of 50 to 500^31^ for homogeneity, however, this may lead to decreased statistical power. In our study, we included all samples and addressed this issue by clustering the samples based on DNA sources using the Gaussian mixture model.

We also developed a novel mtDNA-CN analysis method which corrected for sequencing bias such as GC bias. We benchmarked this method but found that it did not significantly enhance the association findings. Mitochondria have a very narrow range of GC content variation which spans the optimal read capture part of the GC distribution, at least in humans and we suggest that this is why this method did not result in a statistically significant improvement in association findings.

Another challenge of mtDNA-CN analysis is the lack of cell composition data from the same DNA source samples. Gupta et al. demonstrated that blood cell composition influences blood bulk mtDNA-CN^9^, a phenomenon that extends to other tissues. For example, mtDNA-CN varies across different brain regions due to both distinct cellular composition and function. This may partially explain why Pyle et al. observed the association between mtDNA-CN and PD only in the substantia nigra but not in the frontal cortex^6^. We demonstrated that it is feasible to use cell composition data in cohort analysis employing two approaches: (i) using RNA-seq to estimate cell proportions, which is valid under the assumption that RNA and DNA were extracted from the same sample, and (ii) using direct measurements of blood cell type composition using a standard platform.

Of note Yang et al. estimated cell type composition in RNA-seq using xCell^32^, but this method produces enrichment scores rather than percentages, rendering it unsuitable for cell composition correction to be used in mtDNA-CN analysis. Instead, we estimated cell-type proportions using CIBERSORTx^21^ and validated its performance using a test dataset with ground truth cell proportions.

Our findings underscore the importance of analyzing cohorts with cell composition data. It is imperative to ensure that this data is collected at the same time as when performing DNA extraction for the purpose of mtDNA-CN analysis. as cell composition appears to be the main driver of PD associations with mtDNA-CN in this study and cell composition furthermore changes over time. We hypothesize that this will also have confounded previously published studies where no such corrections were performed, usually because the additional data required was missing. This suggests that all studies that intend to work with mtDNA should endeavour to generate and provide cell composition, using the same sample, with the preferred approach being to use the Beckman Coulter method.

Both cell composition correction methods rely on mathematical methods to estimate DNA sources and cell compositions. The RNA-seq based method is likely to be the less accurate method. CIBERSORTx, the deconvolution method we employed, like all signature gene-based methods, relies heavily on reference gene expression data, which may not fully represent the complexity of all possible cell types and states. Also, it may not always distinguish between closely related cell subtypes with similar gene expression profiles and artificially deflate variation in estimates. This lack of resolution can limit the biological insights gained from the RNA-seq deconvolution results. This could explain why there is still a significant association between mtDNA-CN and PD after cell composition correction in the discovery study but not in the replication study.

In summary, our findings indicate that blood mtDNA-CN is not a biomarker of mitochondrial dysfunction for PD but confounds a potential immune signature which we were able to identify instead and which merits further investigation. We arrived at our conclusions using two of the largest datasets in the world, leveraging very recent findings^9^.

## Methods

### Data Cohorts

#### AMP PD

The AMP PD (https://amp-pd.org/) is a collaborative research initiative aimed at advancing the understanding and treatment of PD. The dataset comprises diverse and extensive information from individuals with PD, including clinical, genetic, and biomarker data. Participants contribute detailed clinical histories, demographic information, and undergo various assessments, such as cognitive and motor function evaluations. Genetic data, obtained through WGS, provides insights into the genetic foundations of PD, while biomarker data, including neuroimaging and biofluid analyses, offer valuable insights into disease progression. Macrogen and the Uniformed Services University of Health Sciences (USUHS) conducted all sequencing using the Illumina HiSeq XTen sequencer. Data were aligned to the GRCh38 reference genome. Access to the AMP PD tier two data, including genetic information, was obtained through the application process. All the individual-level analyses were performed on the Terra platform.

#### UKB

The UKB is a major biomedical database that aggregates data from approximately 500,000 participants aged 40 to 69 in the United Kingdom^33^. This extensive dataset encompasses comprehensive information on genetic, clinical, and lifestyle details. In this study, individuals with PD were identified from hospital episode diagnosis, primary care records, and death registries (fields 41234, 42040, and 40023), using all coding that mapped to ICD10 G20 (Parkinson’s disease) in the UKB’s code mapping tables (Resource 592). The first round of WGS, released in late 2021 (Batch 1), involved selecting about 200,000 samples using a pseudorandom approach to ensure cohort representativeness, with 1,728 participants diagnosed with PD. In late 2023, the second round of WGS (Batch 2) was released for the remaining ∼300,000 samples, including 2,562 participants with PD. DNA samples were extracted from buffy coat obtained from participants^34^. Samples underwent sequencing using Illumina NovaSeq6000 technology by two sequencing providers, deCODE Genetics and the Wellcome Trust Sanger Institute. Data were aligned to GRCh38 before undergoing contamination and data quality control. Access to all UKB data was granted on 18^th^ June 2019, application #36610. mtDNA-CN estimation was carried out on the UKB Research Analysis Platform, DNAnexus.

### mitoCN

The existing mtDNA-CN estimators using WGS data, including mtSwirl, assume that reads are uniformly distributed in their alignment to the reference genome and utilize the following formula^9,11^,

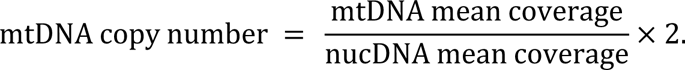

In this study, we introduce mitoCN, a novel method for estimating mtDNA-CN using alignment depth from WGS data while adjusting for coverage bias stemming from homology regions and GC content. mitoCN requires aligned short-read sequencing data in BAM or CRAM format. Aligned reads are filtered out if they have low mapping quality (<30) or SAM alignment flag 3844, which includes: 1) unmapped reads, 2) reads not designated as primary alignment, 3) reads failing platform/vendor quality checks, 4) PCR or optical duplicates, and 5) supplementary alignment. Subsequently, it segments reads into 100-base read bins. To adjust homology bias, we exclusively consider “unique” regions with mappability = 100%. To account for GC bias, we initially selected 100bp read bins with the same GC content range in mtDNA, from 30% to 60%, and then grouped them into 6 clusters with 5% intervals. Hence, we assume that read counts in disjoint read segments are independent and follow the distributions described below:

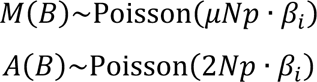

where, *B* denotes a 100bp genomic interval (read bin), and we denote the number of reads aligning into bin *B* on mtDNA as *M*(*B*) and for autosome as *A*(*B*). The parameter *μ* represents the mitochondrial DNA copy number (mtDNA-CN), *Np* denotes the average coverage per read bin, which equals the ratio of the total number of reads to the total number of read bins, and β_*i*_ represents the GC bias parameter in each GC group (*i* = 1,2, …, 6) (Supplementary Notes).

Additionally, to enhance computational efficiency, we employed mosdepth (version 0.2.9) for coverage calculation, a tool that operates nearly twice as fast as the next fastest option, samtools ^35,36^. Moreover, employing the same methodology used for estimating mtDNA-CN, mitoCN enables the assessment of copy numbers for sex chromosomes (chromosomes X and Y). Aberrations in sex chromosome copy numbers, such as XXY and XYY, are not uncommon in the general population and have been linked to specific disorders, such as Klinefelter syndrome and Jacobs syndrome^37,38^.

To compare mitoCN with mtSwirl^9^ (https://github.com/rahulg603/mtSwirl), we applied mtSwirl (v2.5_MongoSwirl_Single) to the AMP PD datasets on the Terra platform. It was observed that the mtSwirl output file (https://github.com/rahulg603/mtSwirl/issues) lacked results for both mtDNA-CN and mean nucDNA coverage. In response, we forked the repository and added commands to compute the mean nucDNA coverage, using samtools idxstats, samtools flagstat, and GATK CollectQualityYieldMetrics^36^. Subsequently, we determined mtDNA-CN using the formula: 2 × mean mtDNA coverage / mean nucDNA coverage. To compare the estimates from mtSwirl and mitoCN, we measured concordance using the square of the correlation (R^2^) between the two estimates and the percent mtDNA-CN change with mtSwirl using the following formula:

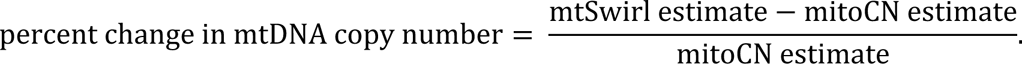

### Statistical Analysis

For cohort association tests, we used robust linear models to mitigate the impact of outliers on regression estimates, utilizing the “rlm()” function from the MASS R package (version 7.3-60)^39,40^. Log-transformed mtDNA-CN, represented as log(mt), was used as the dependent variable and adjusted for ancestry background using the first 5 principal components (PC1-5). Specifically, the following models were utilized for tests involving covariates (age and sex) and PD-related variables (e.g., diagnosis), respectively:

1. Model for covariates: log(mt) ∼ age + PC1-5; log(mt) ∼ sex + PC1-5.
2. Model for PD-related variables: log(mt) ∼ PD diagnosis + age + sex + PC1-5.

For meta-analysis, random-effect (RE) meta-analysis models were applied, fitted with restricted maximum likelihood (REML) estimation using the metafor R package (version 4.4-0)^41^. Two-sample t-tests with unequal variances were employed to assess the significance of the mean difference in binary phenotypes. The p-values, except for those in the meta-analyses, were adjusted to control the FDR at 5% using the Benjamini-Hochberg procedure. All statistical analyses were conducted using R version 4.3.1.

### Blood composition estimation and correction

Whole blood bulk RNA-seq data were obtained from a subset of participants from the PPMI cohort. The gene-level counts from an RNA-seq experiment, featureCounts, were accessed through the Terra platform and were converted to counts per million (CPM) using the edgeR R package (version 4.0.5)^42,43^. The Ensembl gene IDs from the featureCounts matrix were annotated using the biomaRt R package (version 2.58.0)^44,45^ to retrieve associated HUGO gene symbols.

We estimated the cell type proportions from the bulk whole blood RNA-seq data using CIBERSORTx^22^. The reference gene expression profile LM22 served as the signature matrix, consisting of 547 genes that differentiate among 22 human hematopoietic cell phenotypes^23^. Notably, as LM22 includes genes only for PBMCs and not platelets, we employed the absolute score for cluster 2 to reflect the absolute proportion of each cell type in a mixture. B-mode (bulk mode) batch correction was applied to address technical differences between the LM22 signature matrix derived from microarrays and the input bulk RNA-seq data. Significance analysis was conducted with 100 permutations. The absolute score, reflecting the absolute proportion of each cell type in a mixture, was utilized.

To assess the performance of the CIBERSORTx and validate the cell composition estimation, we employed a validation cohort comprising whole blood samples from 12 healthy adults, sourced from the CIBERSORTx website (https://cibersortx.stanford.edu/download.php). This cohort offers ground truth cell proportions determined by direct flow cytometry and whole blood bulk RNA-seq data. Applying the same parameters described above, we estimated the cell composition of the validation cohort and compared the estimates with the ground truth proportions using Pearson correlation tests. Despite systematically over- or under-estimates of some cell types, such as neutrophils and monocytes, the overall proportion estimates show significantly positive correlations with the true measurements (R: 0.65-0.95, Supplementary Fig. 19A). Given our intention to use the cell composition for covariate adjustment, the relative values across samples are crucial, and the estimation bias will not impact the downstream analyses. Boxplots illustrating the range of proportion estimates from the validation cohort and healthy controls in the AMP PD data demonstrate a consistent distribution across cell types (Supplementary Fig. 19B-C). These results suggest that the computationally estimated proportions are reliable.

We utilized total absolute score as the proportion of white blood cells in a mixture of whole blood and selected 8 of 22 cell types with proportions >0.01. These include naïve B cells, naïve CD4 T cells, resting memory CD4 T cells, activated memory CD4 T cells, resting NK cells, monocytes, resting mast cells, and neutrophils. Using a stepwise model selection procedure we excluded three blood variables: activated memory CD4 T cells, resting NK cells, and monocytes. Adjusted mtDNA-CN was defined using the residuals of the following model: log(mtDNA-CN) ∼ naïve B cells + naïve CD4 T cells + resting memory CD4 T cells + resting mast cells + neutrophils + white blood cells.

### Polygenic risk score

Using the summary statistics made available by Gupta et al. 2023^9^ from the across-ancestry meta-GWAS (Supplementary Table 1C) we calculated the PRSs for both raw and adjusted mtDNA-CN for all individuals in the AMP PD v3 dataset with available WGS data. The GWAS for adjusted mtDNA-CN identified 92 Linkage Disequilibrium (LD)-independent signals, with 88 variants present in the AMP PD dataset. One missing variant, rs578069621, was replaced with SNP in perfect LD (D’ = 1), rs141447648, using the LDProxy^46^ tool. The resulting 89 SNP PRS was calculated using the --score flag in PLINK 2.0^47^. In the case of raw mtDNA-CN, a similar approach was employed, leading to the utilization of 134 out of 141 variants for PRS calculation. The two PRSs are in positive correlation (R = 0.62, *p* < 2.2e-16).

To validate our calculation, we demonstrated a negative correlation between PRSs and kinship coefficients (Supplementary Fig. 20), suggesting that individuals with familial relationships exhibit more similarity in their PRSs compared to unrelated individuals. Additionally, we show that both mtDNA-CN PRSs are positively correlated with actual mtDNA-CN estimates from blood samples (Supplementary Table 1D, Supplementary Fig. 9). To investigate the associations between PD and the mtDNA-CN PRSs, density plots, and t-tests were utilized to compare the PRSs between healthy controls and individuals diagnosed with PD.

### Bidirectional MR between mtDNA-CN and PD

We performed a bidirectional two-sample MR analysis, employing SNPs as IVs based on summary statistics from GWASs. The summary-level data for PD risk was obtained from a recent meta-analysis of GWASs in the European population^48^. For blood-derived mtDNA-CN, we utilized three GWAS datasets employing different estimation methods, including estimates from WGS with and without adjusting for cell composition^9^, estimates from genotyping data^10^, and estimates from a combination of WES and genotyping arrays^49^. All mtDNA-CN GWAS studies were conducted using UKB data, and the majority of participants had European ancestry (Supplementary Table 1C).

To ensure the selection of valid SNPs as IVs for our study, several criteria were applied. This included filtering SNPs based on p-value thresholds (*p* < 5 × 10⁻⁶ for PD and *p* < 5 × 10⁻⁸ for mtDNA-CN phenotypes), conducting LD clumping (r² = 0.001 with a window size of 10,000 kb) with the “clump_data()” function in the TwoSampleMR R package, aligning the effect alleles of the exposure and outcome variables to the forward strand, and excluding palindromic SNPs.

For the causal estimate, we employed multiple methods, including IVW, MR Egger, weighted median, and weighted mode. To assess the robustness of the causal estimates, sensitivity analyses were conducted, incorporating the heterogeneity test measured by Cochran’s Q statistic and pleiotropy by the MR-Egger intercept test. Furthermore, to evaluate the potential impact of each SNP on the IVW estimate, leave-one-out analyses were performed, systematically removing one SNP at a time. Funnel plots were used to visualize the selection bias of IVs. All statistical analyses were conducted in R software (version 4.3.1) using the R package TwoSampleMR (version 0.5.7)^50,51^.

## Data availability

Access to the AMP PD data is available through the Terra platform upon completion of an AMP PD access application (https://www.amp-pd.org/register-for-amp-pd). UKB phenotype and WGS data can be obtained through the UKB Research Analysis Platform following the submission of a UKB access application (https://ukbiobank.dnanexus.com/landing). Individual-level data, mtDNA-CN estimates, generated as part of AMP PD have been returned to enable utilization of the full individual-level data by the broader scientific community through the Terra workspace (https://app.terra.bio/#workspaces/bahlo_lab_amp_pd/MJFF-021399/data).

## Code availability

The mitoCN software and the code for all analyses are accessible to the public at https://github.com/bahlolab/mitoCN/tree/main/scripts.

## Acknowledgments

We are grateful to Grant Dewson and Tahnee Saunders for their assistance with this study. This work was supported by the Michael J Fox Foundation for Parkinson’s Research (MJFF) and the Shake It Up Australia Foundation (MJFF-021399). MB was supported by an NHMRC Investigator Grant (GNT1195236). This work was also supported by the Australian State of Victoria’s Government’s Operational Infrastructure Support Program, the NHMRC Independent Research Institute Infrastructure Support Scheme (IRIISS), and the Felton Bequest.

Data used in the preparation of the discovery study were obtained from the AMP PD Knowledge Platform. For up-to-date information on the study, visit https://www.amp-pd.org. The AMP PD program is a public-private partnership managed by the Foundation for the National Institutes of Health (NIH) and funded by the National Institute of Neurological Disorders and Stroke (NINDS) in partnership with the Aligning Science Across Parkinson’s (ASAP) initiative; Celgene Corporation, a subsidiary of Bristol-Myers Squibb Company; GlaxoSmithKline plc (GSK); The MJFF; Pfizer Inc.; AbbVie Inc.; Sanofi US Services Inc.; and Verily Life Sciences. ACCELERATING MEDICINES PARTNERSHIP and AMP are registered service marks of the U.S. Department of Health and Human Services.

Clinical data used in the discovery study were obtained from:

i. BioFIND (https://www.michaeljfox.org/news/biofind), sponsored by the MJFF with support from the NINDS;
ii. HBS (https://www.bwhparkinsoncenter.org/biobank/), a collaboration of HBS investigators (full list of HBS investigators found at https://www.bwhparkinsoncenter.org/biobank/) and funded through philanthropy and NIH and Non-NIH funding sources;
iii. PDBP (https://pdbp.ninds.nih.gov/), supported by the NINDS at the NIH (A full list of PDBP investigators can be found at https://pdbp.ninds.nih.gov/policy);
iv. PPMI (https://www.ppmi-info.org/), sponsored by the MJFF and supported by a consortium of scientific partners (list the full names of all the PPMI funding partners found at https://www.ppmi-info.org/about-ppmi/who-we-are/study-sponsors);
v. STEADY-PD3 (https://clinicaltrials.gov/ct2/show/study/NCT02168842) and (vi) SURE-PD3 (https://clinicaltrials.gov/ct2/show/NCT02642393), both funded by the NINDS at the NIH with support from the MJFF.

The Investigators of the six studies have not participated in reviewing the data analysis or content of the manuscript. For up-to-date information on the studies, visit the websites provided above.

The data used in the replication study are obtained from UKB Resource (https://www.ukbiobank.ac.uk/) under Application Number 36610. This work uses data provided by patients and collected by the NHS as part of their care and support.

We would like to thank the participants and their families, without whom these studies would not have been possible.

## Author information

### Contributions

M.B. and L.W. conceived and designed the research. L.W. wrote the manuscript. L.G.F. and M.M. set up pipelines for cloud computing. L.W. and L.G.F. performed individual-level analysis on cloud computing platforms. L.W., L.G.F., J.R., and H.R. performed metadata extraction, generation, and quality control. L.W. performed the discovery analysis. J.H. performed the replication analysis and wrote the replication study section in the manuscript. T.P.S. consulted on the statistical methods. L.W. developed the bioinformatics software with assistance from L.G.F. and M.M. Z.G., T.L., and S.M. provided advice on interpreting the results and assisted in revising the manuscript. M.B. oversaw the study and provided direction, funding, and resources. All authors read and approved the final manuscript.

### Ethics declarations

This study was approved by the Walter and Eliza Hall Institute of Medical Research (WEHI), Human Research Ethics Committee (HREC reference 17/09LR and 22/19). All components of this study were conducted in accordance with the principles embodied within the Declaration of Helsinki.

## Supplementary information

Supplementary Data

Supplementary Table

