## Supplemental Data for "Unveiling Peripheral Immune Dysfunction in Parkinson’s Disease through Analysis of Blood-based Mitochondrial DNA Copy Number"

### Supplementary Notes

#### mitoCN

##### Homology Bias Adjustment

We generated bin annotation data with a bin size of 100 base pairs using the "createBins()" function. This data encompasses chromosome names, the positions of the first and last base pair within each bin, the percentage of characterized nucleotides (A, C, G, or T, i.e. non-N), and the GC content (percentage of C and G nucleotides among non-N nucleotides). Subsequently, we computed the average mappabilities per read bin using the "calculateMappability()" function. This involved employing a mappability file in bigWig format and the bigWigAverageOverBed binary. Specifically, we derived a 50-mer mappability file with 2 mismatches using GenMap (v1.3.0)[1] for the hg38 genome assembly. For hg19, we utilized the "wgEncodeCrgMapabilityAlign50mer.bigWig" file from the ENCODE's download section of the UCSC Genome Browser. The bigWigAverageOverBed binary can be downloaded from the UCSC Genome Browser's Other Utilities section. Additionally, we determined the percentage overlap between the generated bins and ENCODE's Blacklisted Regions via the "calculateBlacklist()" function. Any read bins with mappability below 100% or overlap with blacklist regions exceeding 0% were excluded. These analyses were performed using R software (version 4.3.1) with the QDNAseq[2] R package (version 1.38.0).

##### GC Bias Adjustment

After selecting read bins with mappability = 100%, the GC content range on mtDNA spans from 30% to 60%. We segmented this range into six groups, each with 5% intervals, denoted as $g_i$ ($i$ = 1, 2, …, 6). Let $\mathcal{M}_i$ and $\mathcal{A}_i$ represent the collection of 100bp read bins $B$, we use in the mitochondrial and autosomal genomes, respectively, falling within the GC content range $g_i$. Write $|\mathcal{M}_i| = m_i$ and $|\mathcal{A}_i| = a_i$ for the number of read bins in these collections. We use $m_+ = \sum_{i=1}^{6} m_i = 20$ mitochondrial bins with differing GC content, all with 100% mappability. We then randomly select 20 bins in each chromosome that match the characteristics of the mitochondrial bins in terms of GC content and mappability, repeating this process $k$ times. Using bins from only one autosome, $a_i = km_i$ so $a_+ = km_+ = 20k$, while using bins from all 22 autosomes, $a_+ = 440k$.

$M(B)$ is the number of reads with 3' end in the mitochondrial bin $B$ while $A(B)$ is the number of reads with 3' end in the autosomal bin $B$. Assume that for $B \in \mathcal{M}_i$, we have $M(B) \sim Poisson(\mu N p \beta_i)$, while for $B \in \mathcal{A}_i$, we assume that $A(B) \sim Poisson(2Np\beta_i)$.

The log-likelihood generated by the bin counts under these assumptions is,

$$l = \Sigma_i \left[\Sigma_{B\in\mathcal{M}_i} \{-\mu N p \beta_i + M(B) log\,(\mu N p \beta_i)\} + \Sigma_{B\in\mathcal{A}_i} \{-2Np\beta_i + A(B) log\,(2Np\beta_i)\}\right]$$

which when we write $M_i = \Sigma_{B\in\mathcal{M}_i} M(B)$ and $A_i = \Sigma_{B\in\mathcal{A}_i} A(B)$, simplifies to

$$l = \Sigma_i \left[-\mu Npm_i\beta_i + M_i log(\mu Np\beta_i) - 2Npa_i\beta_i + A_i log(2Np\beta_i)\right].$$

We differentiate this with respect to $\mu$ and the $\{\beta_i\}$ and equate the derivatives to 0, getting estimating equations for

$$\mu = \frac{\Sigma_i \Sigma_{B\in\mathcal{M}_i} M(B)}{Np\Sigma_i m_i\beta_i} = \frac{M}{Np\Sigma_i m_i\beta_i} \ldots\ldots (1)$$

$$\beta_i = \frac{T_i}{Np(\mu m_i + 2a_i)} \ldots\ldots\ldots\ldots\ldots (2)$$

where $M$ is the total number of reads mapping to mtDNA, $T_i = \Sigma_{B\in\mathcal{M}_i} M(B) + \Sigma_{B\in\mathcal{A}_i} A(B)$.

#### Correction in the Gupta et al., 2023

We have identified an error in the Gupta et al., 2023 study[3], where the effect sizes of the genome-wide association studies (GWASs) were incorrectly signed for the reference allele listed. Consequently, this error has impacted several interpretations in the paper, such as Figure 1E, and the direction of all Mendelian randomization (MR) plots (Figure 1G, 1H, EDF4G, EDF4H, and EDF6). We have communicated our concerns to the authors, and they have addressed this issue by updating the GWAS Catalog summary statistics on April 5, 2024, by swapping the allele listed in the "effect allele" and have submitted a corresponding correction to Nature. The GWAS summary statistics utilized in this study were those post-correction.

Furthermore, it's important to note that the output file from the mtSwirl v2.5_MongoSwirl_Single pipeline lacks results for both mitochondrial DNA copy number (mtDNA-CN) and mean nuclear DNA (nucDNA) coverage (https://github.com/rahulg603/mtSwirl/issues). Users are required to modify the pipeline by adding commands for nucDNA calculation before applying it on Terra and calculate mtDNA-CN independently. This may impact the reproducibility of the tool. In contrast, mitoCN is a single-command tool and is more user-friendly.

### Supplementary Figures

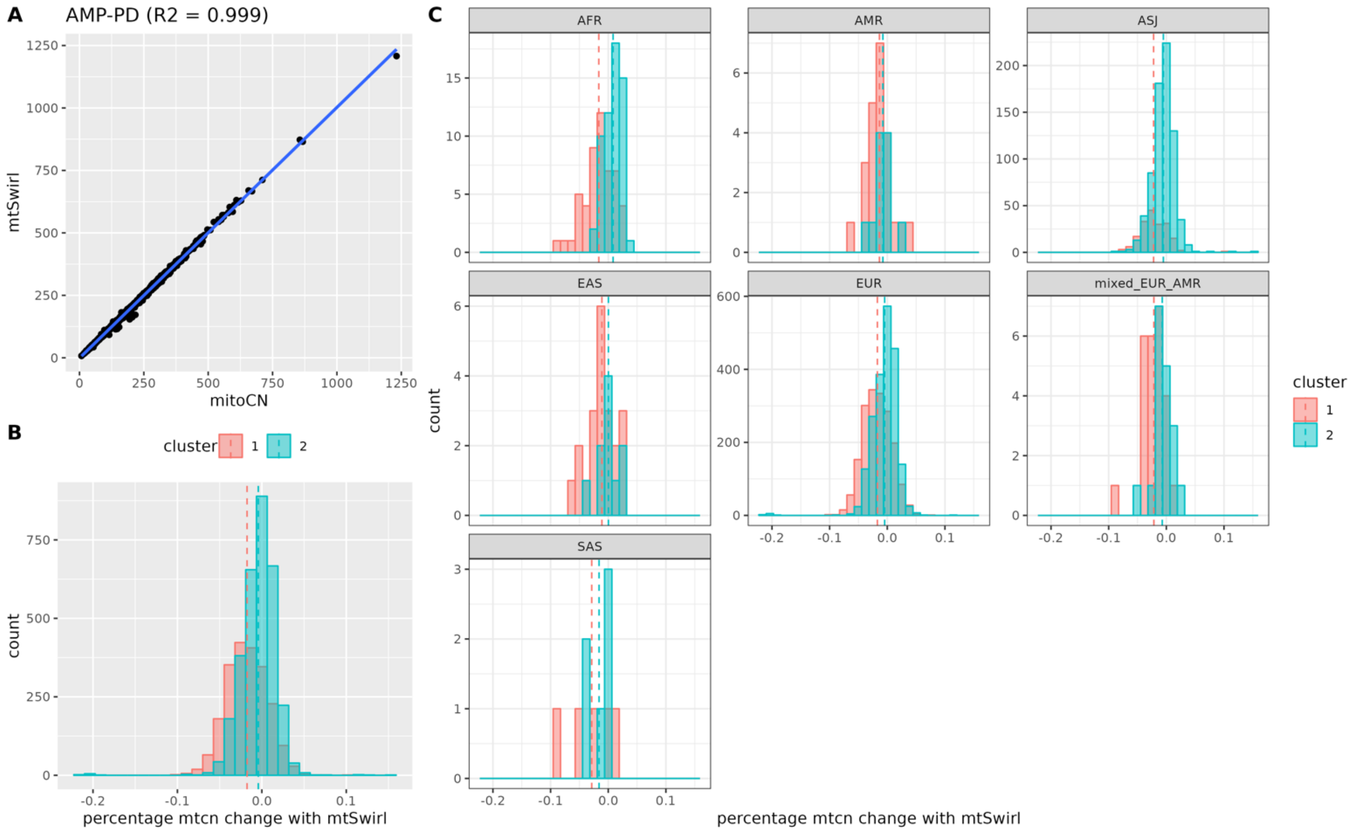


**Supplementary Fig. 1 Comparison of estimates from mtSwirl and mitoCN.**

**a**, scatter plot illustrates a high concordance in mtDNA-CN estimates between mitoCN and mtSwirl ($R^2 = 0.999$, $p$ <2.2e-16); **b**, histogram shows the percent change in mtDNA-CN estimated from mitoCN and mtSwirl. The average percent change for cluster 1 is -1.8%, and for cluster 2, it is -0.4%; **c**, percent change in mtDNA-CN estimated using mitoCN versus mtSwirl, grouped by inferred nuclear ancestry. The superpopulation groups that can be inferred include AFR, AMR, ASJ, EAS, EUR, SAS, for African, American, Ashkenazi Jewish, east Asian, European, and south Asian, respectively. Cluster 1 is referred to as "platelet-depleted blood samples", while cluster 2 is referred to as "platelet-abundant blood samples".

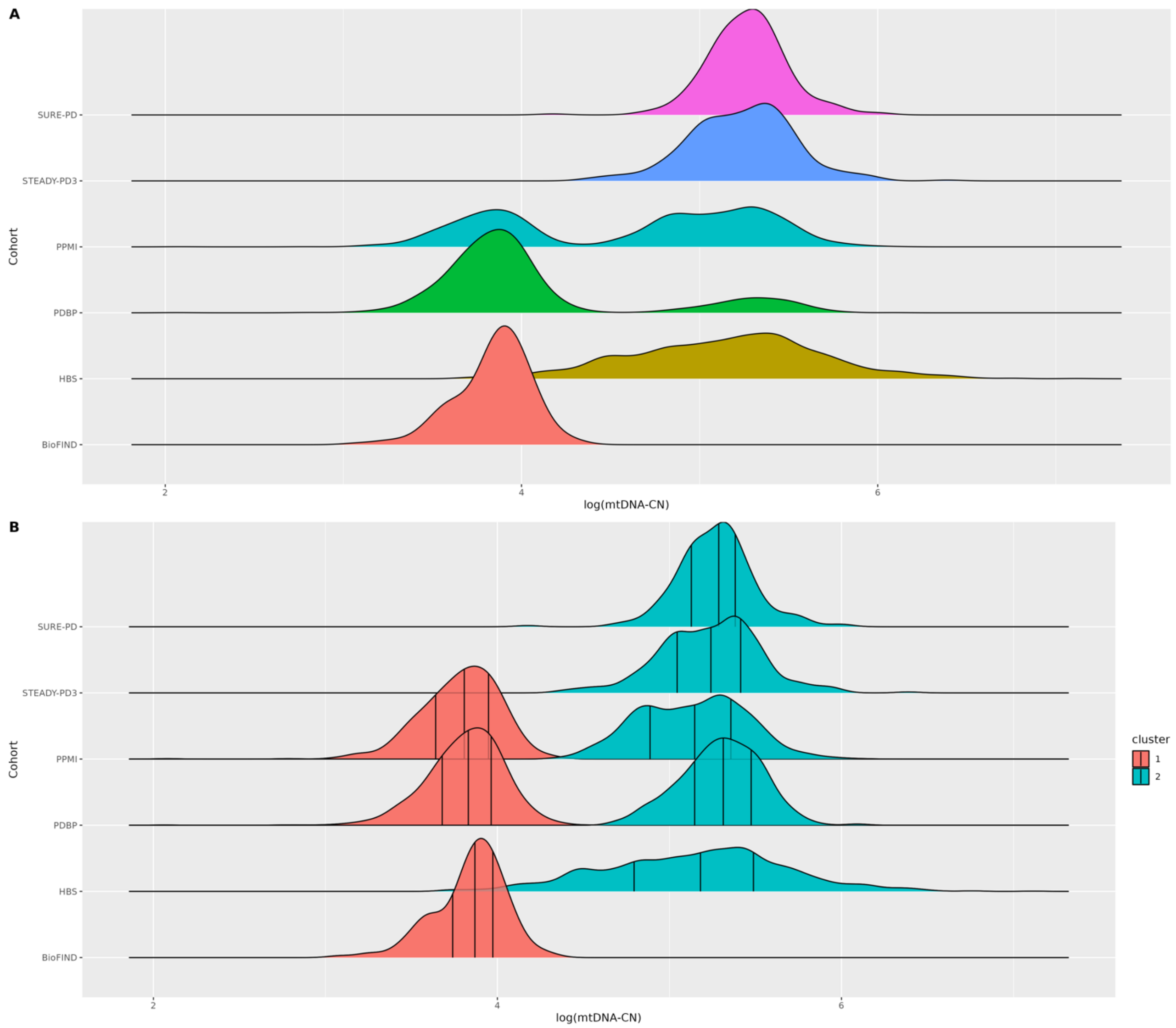


**Supplementary Fig. 2 Log scaled mtDNA-CN distribution in AMP PD cohorts.**

**a,** two distinct distributions in the mtDNA-CN estimates, suggesting that DNA samples were extracted from two types of blood samples; **b**, two clusters classified using Gaussian mixture model.

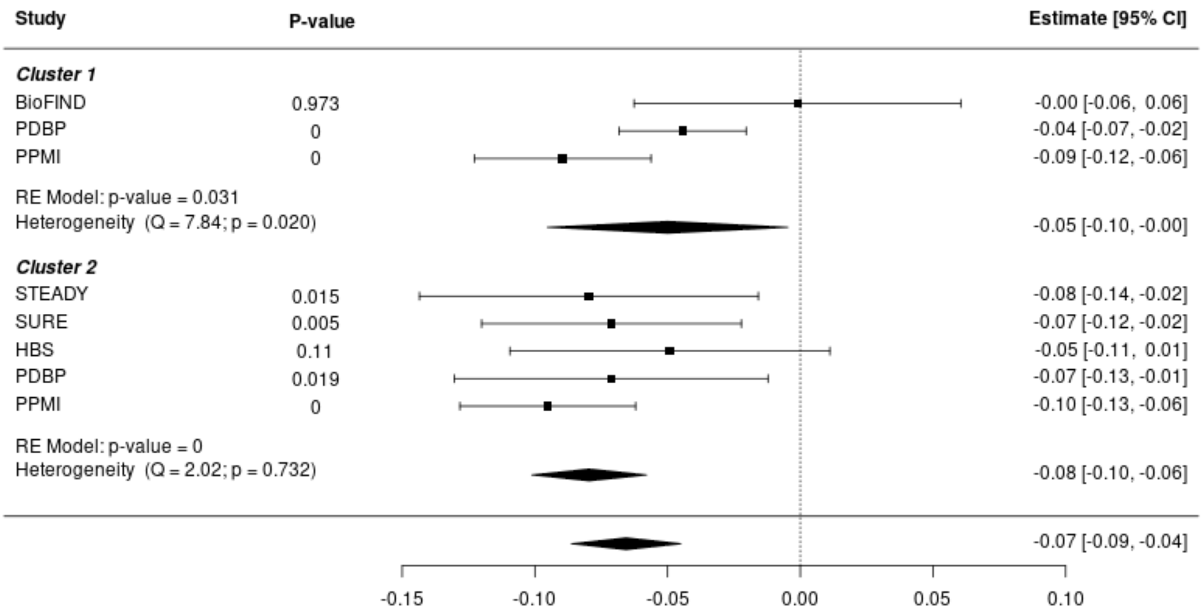


**Supplementary Fig. 3 Meta-analysis of association tests between blood mtDNA-CN and sex.** Both blood sample types confirmed the association between sex and mtDNA-CN, indicating that mtDNA-CN tends to be lower in males compared to females. The absolute effect size estimated from cluster 2 (beta = -0.08, $p$ <0.0001), which comprises platelet-abundant samples, is greater than that of cluster 1 (beta = -0.05, $p$ = 0.03). Additionally, the variability of estimate from cluster 2 (95% CI [-0.09, -0.04]) is smaller than cluster 1 (95% CI [-0.10, 0]).

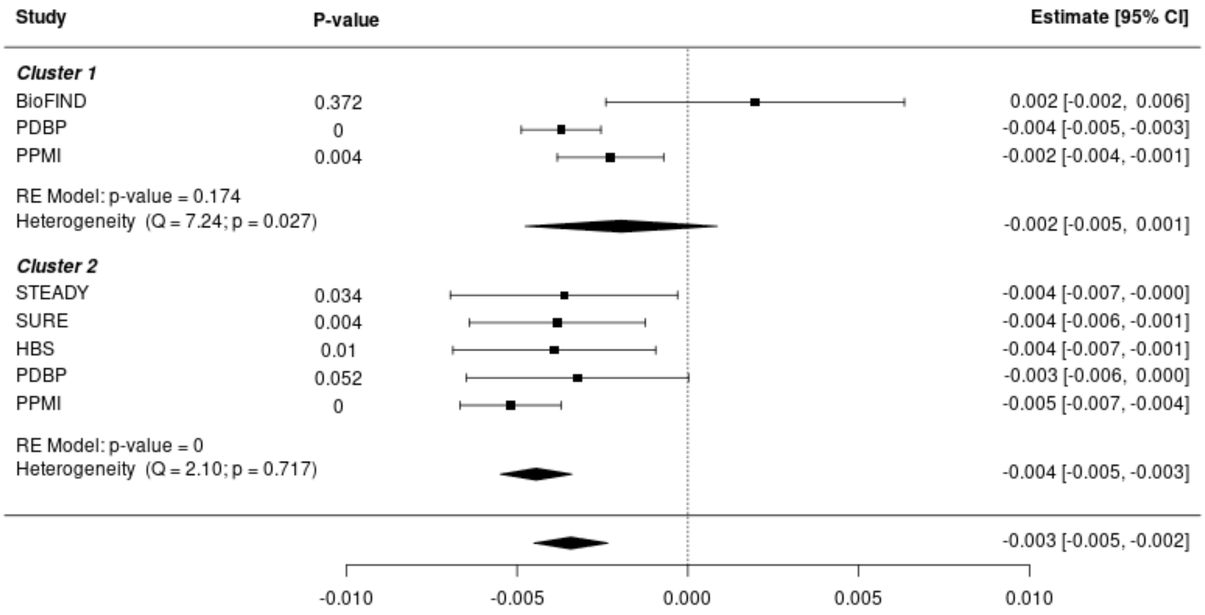


**Supplementary Fig. 4 Meta-analysis of association tests between blood mtDNA-CN and age.** For cluster 2, the meta-analysis results indicate a significant decline in mtDNA-CN with age (beta = -0.004, *p* <0.0001). However, the association is not significant in cluster 1, mainly due to the heterogeneity of the BioFIND cohort. Upon excluding the BioFIND cohort, there is no evidence of heterogeneity between the PPMI and PDBP cohorts (Q = 2.12, *p* = 0.15), and the association becomes significant (beta = -0.003, *p* <0.0001).

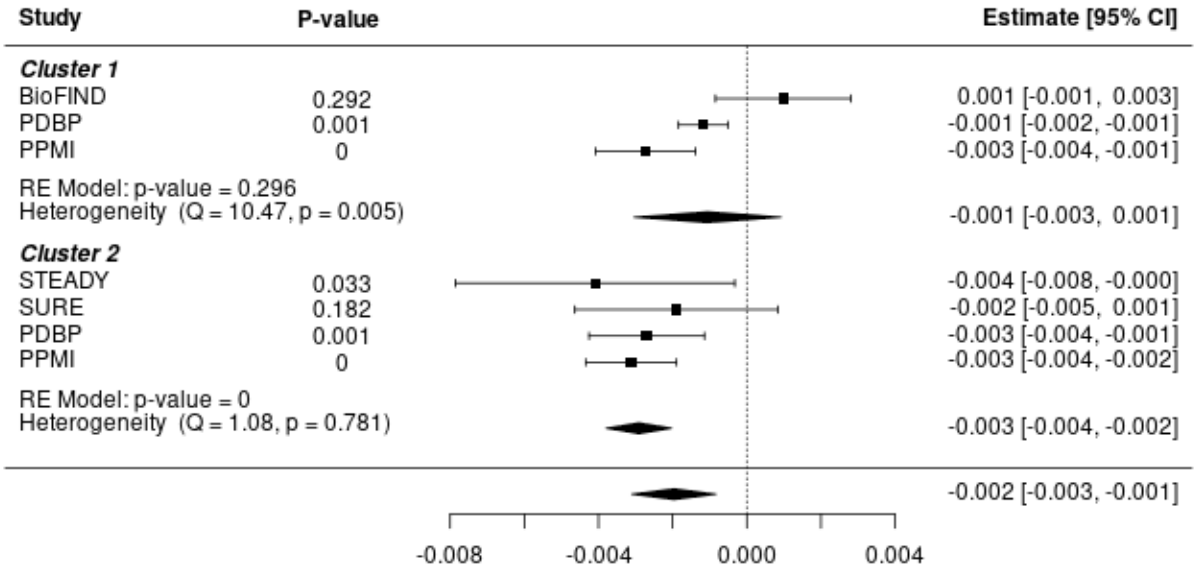


**Supplementary Fig. 5 Meta-analysis of association tests between blood mtDNA-CN and motor examination (MDS-UPDRS III).**

In cluster 2, the meta-analysis results reveal a significant correlation, indicating that lower mtDNA-CN is associated with more severe motor symptoms (beta = -0.003, $p$ <0.0001). However, in cluster 1, this association lacks significance, mainly due to the heterogeneity from the BioFIND cohort. After excluding the BioFIND cohort, no significant heterogeneity is detected between the PPMI and PDBP cohorts (Q = 1.08, $p$ = 0.78), and the association becomes significant (beta = -0.003, $p$ <0.0001).

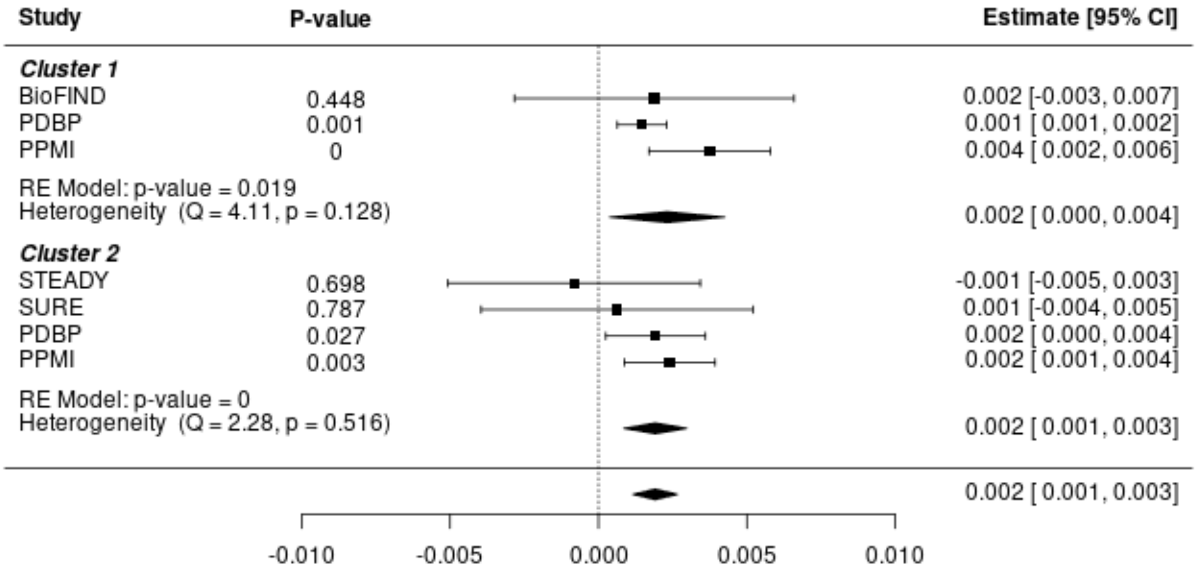


**Supplementary Fig. 6 Meta-analysis of association tests between mtDNA-CN and activities of daily living (ADL).**

Both clusters validate the correlation between ADL and mtDNA-CN, suggesting that reduced levels of mtDNA-CN are linked to impaired performance in daily activities. The effect size estimated from both clusters is 0.002. Moreover, the variability of the estimate from cluster 2 (95% CI [0.001, 0.003]) is narrower compared to cluster 1 (95% CI [0, 0.004]), indicating greater precision, and it is also more statistically significant.

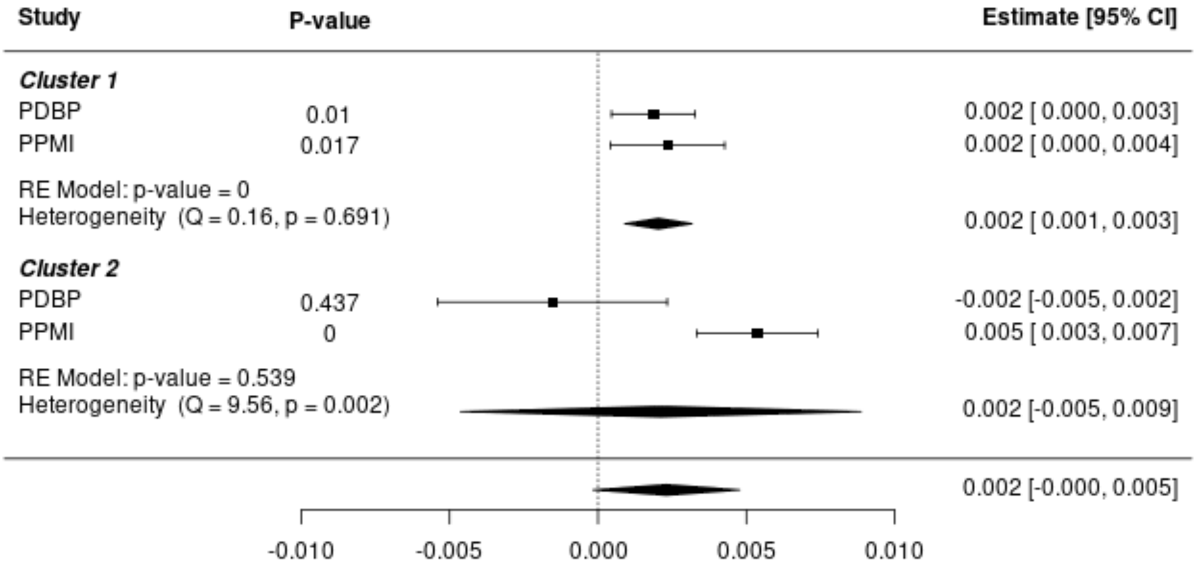


**Supplementary Fig. 7 Meta-analysis of association tests between mtDNA-CN and olfactory impairment (UPSIT).**

The olfactory bulb has been identified as one of the primary regions where PD is thought to initiate, and the UPSIT score has been linked to the severity of PD. In cluster 1, the meta-analysis reveals a significant association between lower mtDNA-CN and olfactory dysfunction (beta = 0.002, $p$ <0.0001). However, in cluster 2, this association lacks significance, likely attributable to the heterogeneity of the PDBP and the PPMI cohorts (Q = 9.56, $p$ = 0.002). Notably, the PDBP cohort in cluster 2 exhibits significant differences from the other cohorts. Focusing solely on the PPMI cohort within cluster 2, we observe a larger effect size compared to cluster 1, possibly due to mtDNA enrichment from platelets.

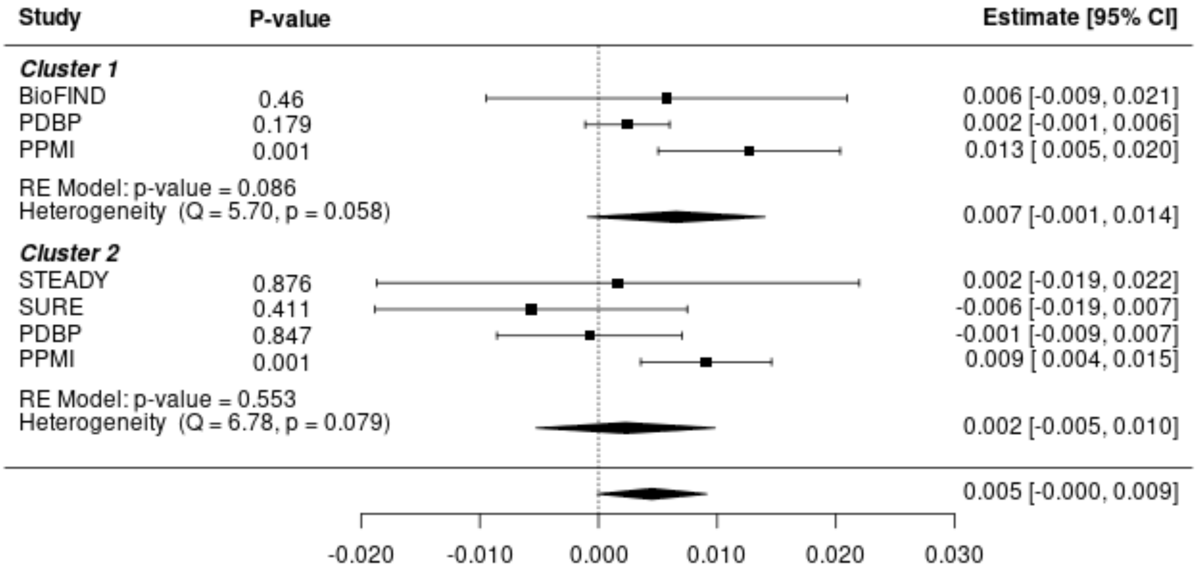


**Supplementary Fig. 8 Meta-analysis of association tests between mtDNA-CN and cognitive impairment (MoCA).**

Through cohort analysis, the association between mtDNA-CN and cognitive impairment was solely identified within the PPMI cohort across both clusters, while not observed in other cohorts. The meta-analysis results from both clusters indicate no significant associations.

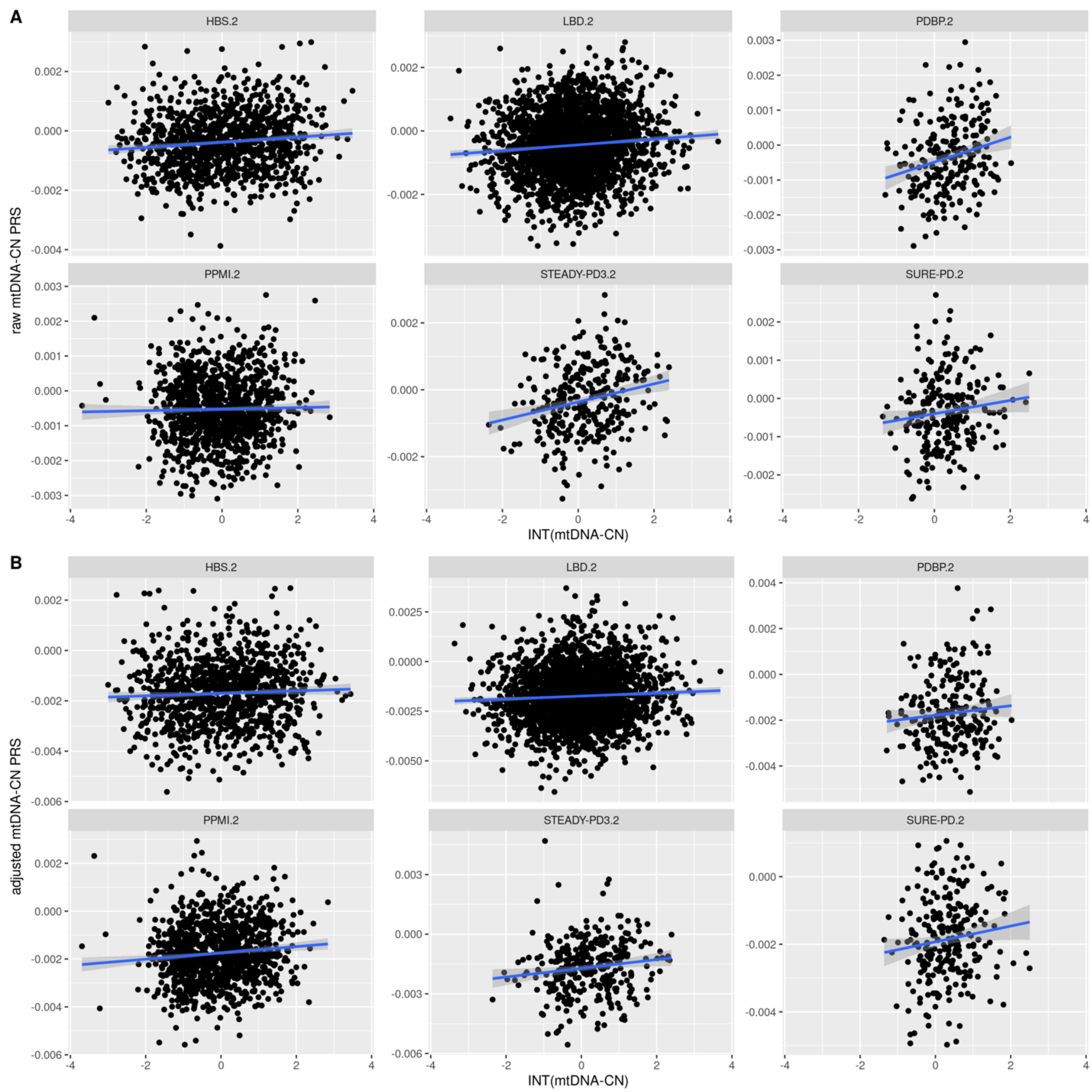


**Supplementary Fig. 9 Correlation analysis between mtDNA-CN PRSs and actual mtDNA-CN estimates from whole blood samples in each cohort.**

**a**, scatter plots illustrate the positive correlation between raw mtDNA-CN PRS and the actual mtDNA-CN from cluster 2 in each cohort; **b**, scatter plots depict the positive correlation between adjusted mtDNA-CN PRS and the actual mtDNA-CN from cluster 2 in each cohort. The mtDNA-CN estimates underwent transformation using a rank-based inverse normal transform (INT).

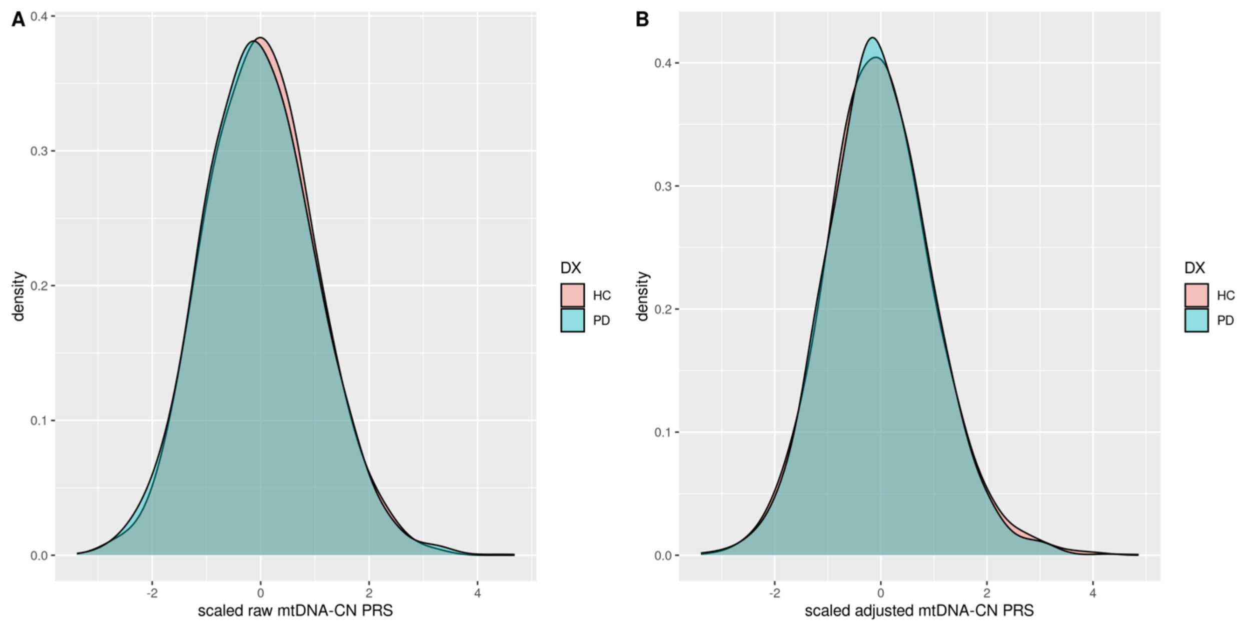


**Supplementary Fig. 10 Density plots of mtDNA-CN PRSs in healthy controls (HC) and individuals diagnosed with PD.**

**a**, scaled raw mtDNA-CN PRS in HC and PD; **b**, scaled adjusted mtDNA-CN PRS in HC and PD.

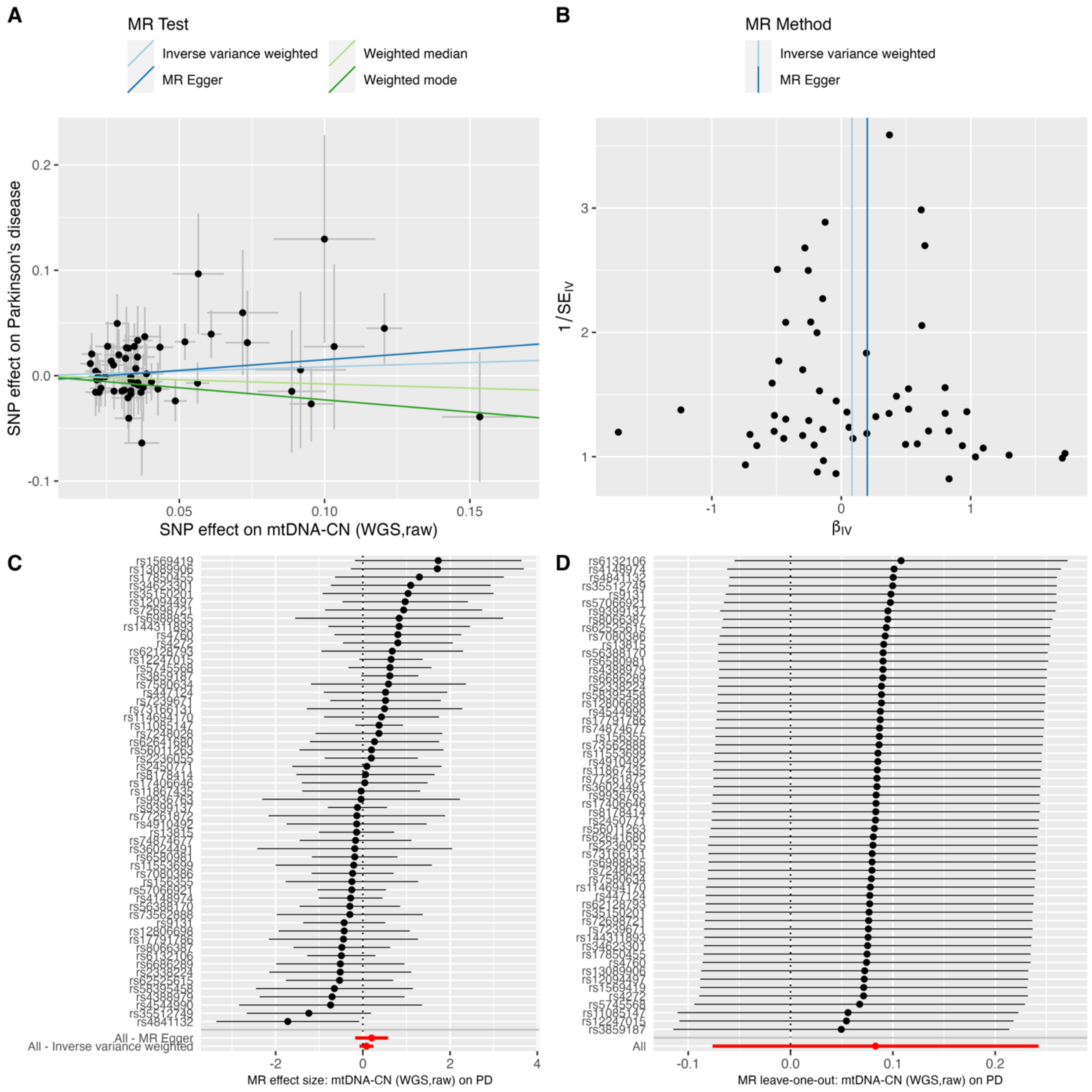


**Supplementary Fig. 11 Mendelian Randomization analysis of mtDNA-CN (WGS, raw) on PD.**

**a**, scatter plots showing the causal association; **b**, funnel plots; **c**, single SNP effect size plot; **d**, leave-one-out plot.

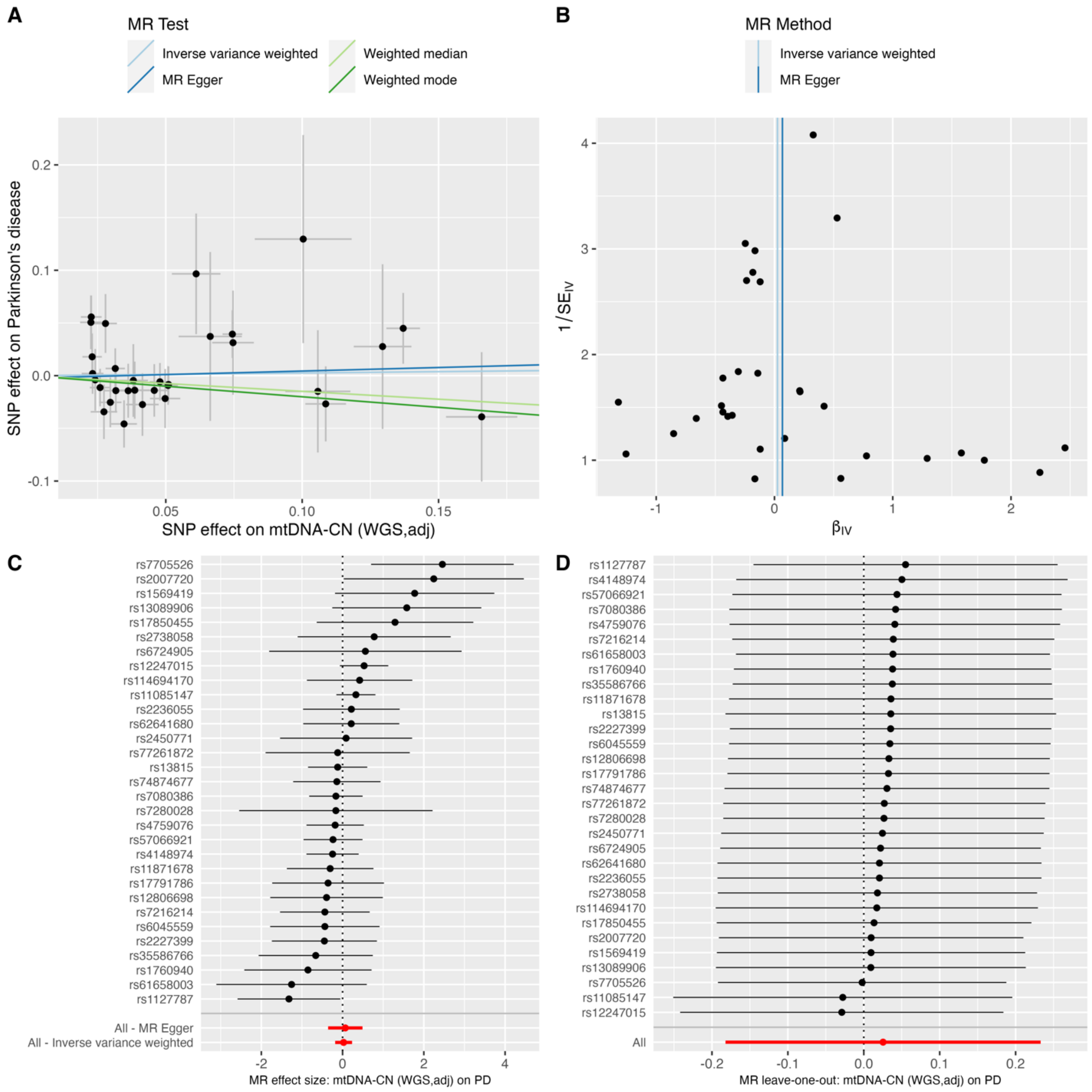


**Supplementary Fig. 12 Mendelian Randomization analysis of mtDNA-CN (WGS, adjusted) on PD.**

**a**, scatter plots showing the causal association; **b**, funnel plots; **c**, single SNP effect size plot; **d**, leave-one-out plot.

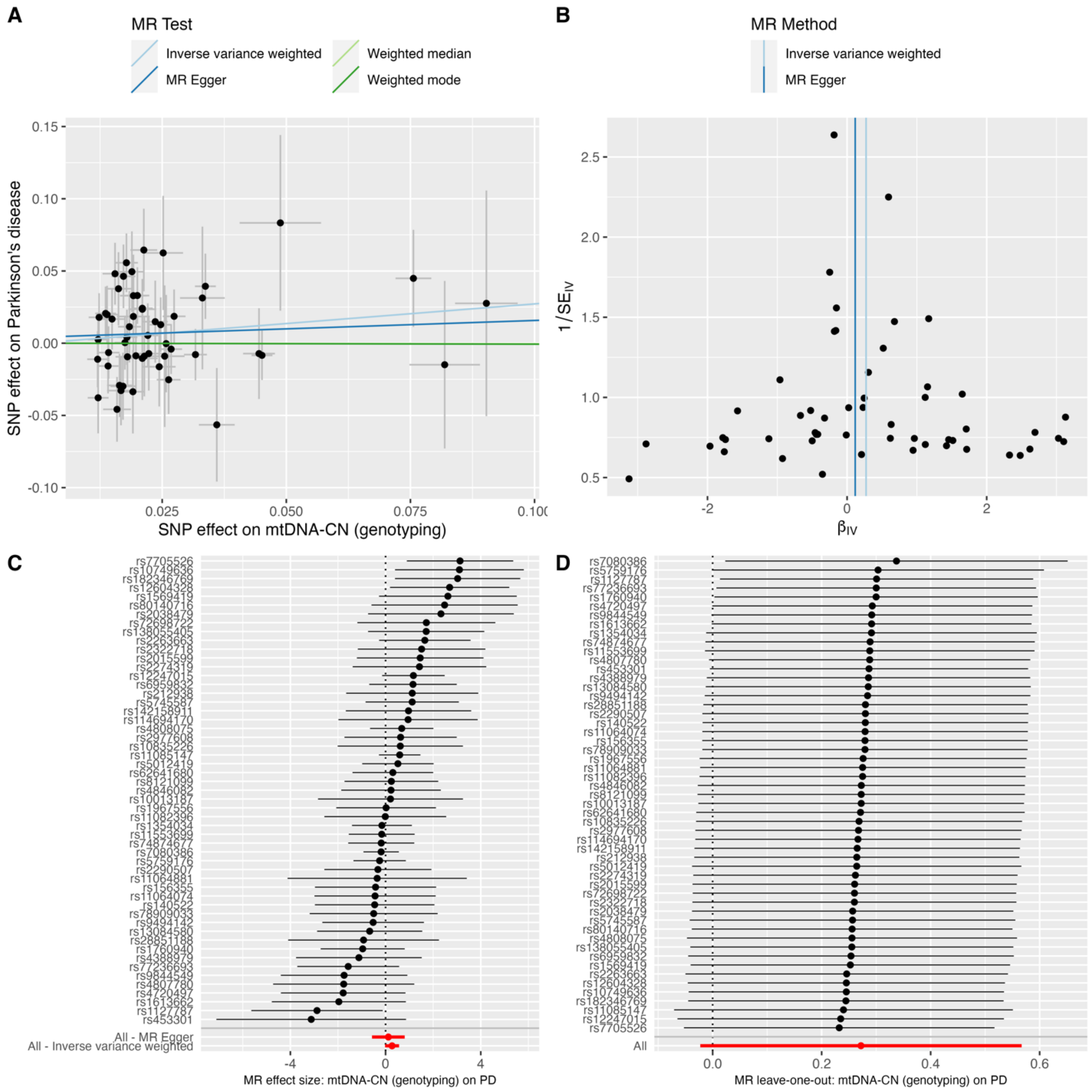


**Supplementary Fig. 13 Mendelian Randomization analysis of mtDNA-CN (genotyping) on PD.**

**a**, scatter plots showing the causal association; **b**, funnel plots; **c**, single SNP effect size plot; **d**, leave-one-out plot.

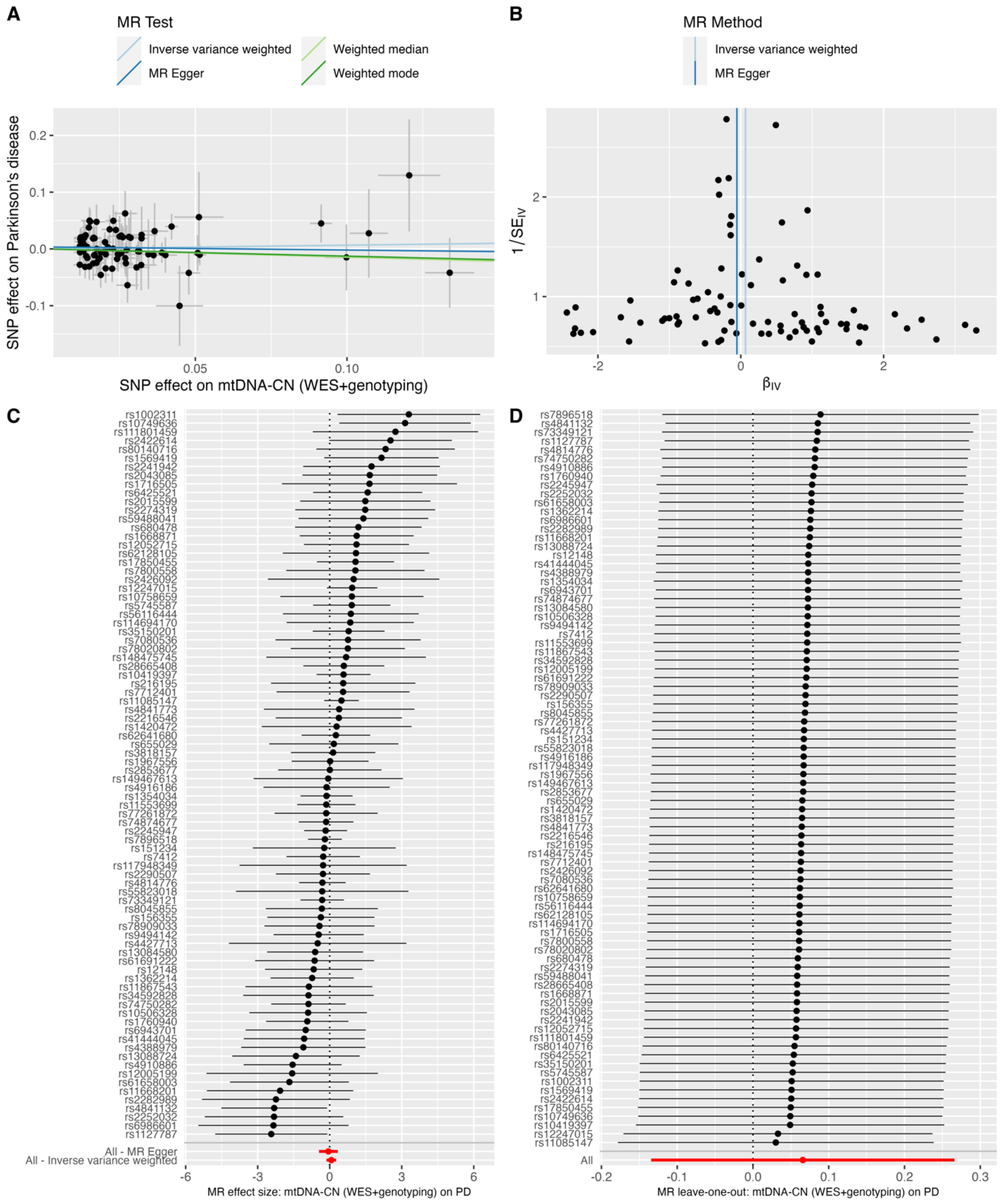


**Supplementary Fig. 14 Mendelian Randomization analysis of mtDNA-CN (WES + genotyping) on PD.**

**a**, scatter plots showing the causal association; **b**, funnel plots; **c**, single SNP effect size plot; **d**, leave-one-out plot.

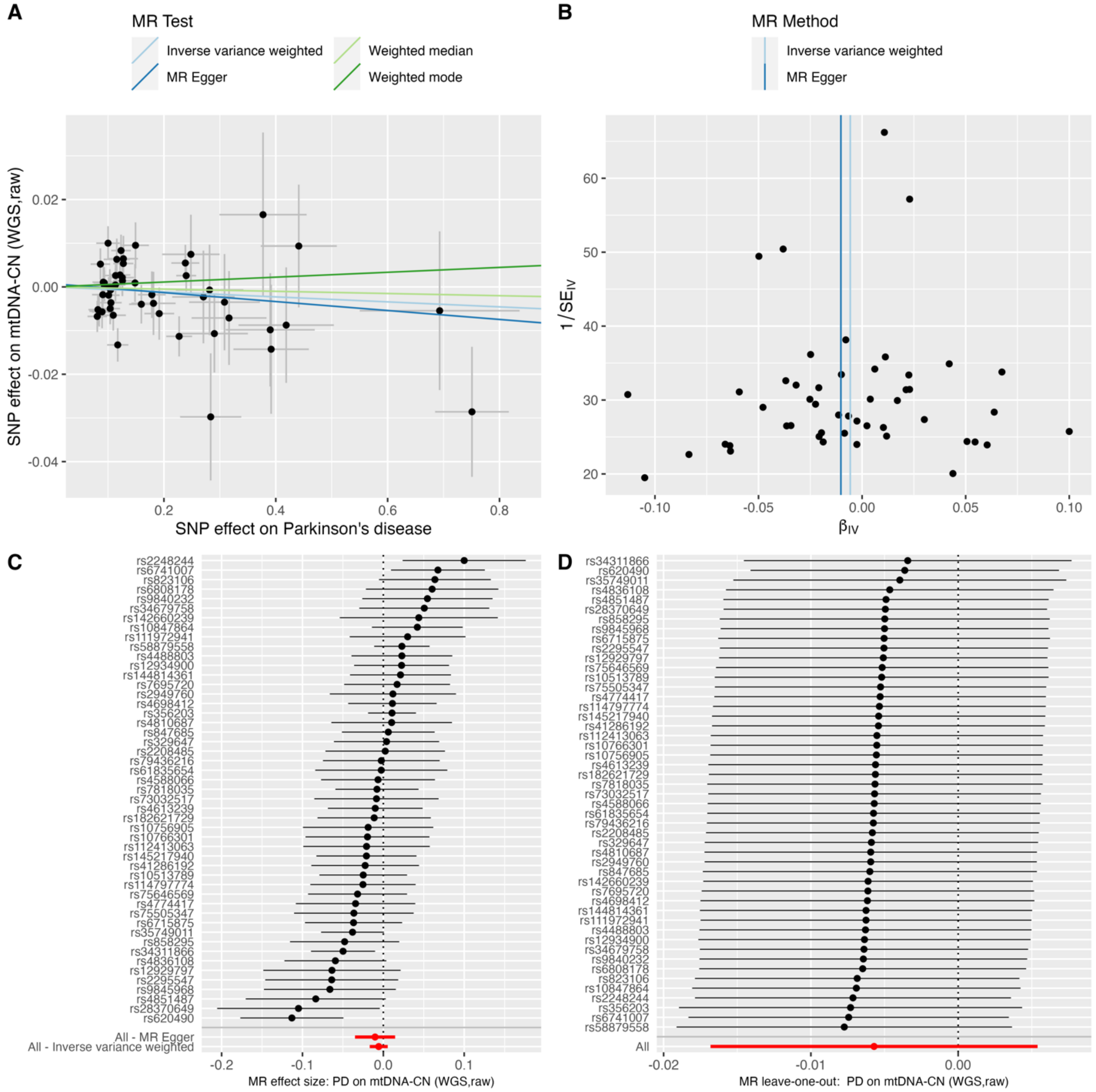


**Supplementary Fig. 15 Mendelian Randomization analysis of PD on mtDNA-CN (WGS, raw).**

**a**, scatter plots showing the causal association; **b**, funnel plots; **c**, single SNP effect size plot; **d**, leave-one-out plot.

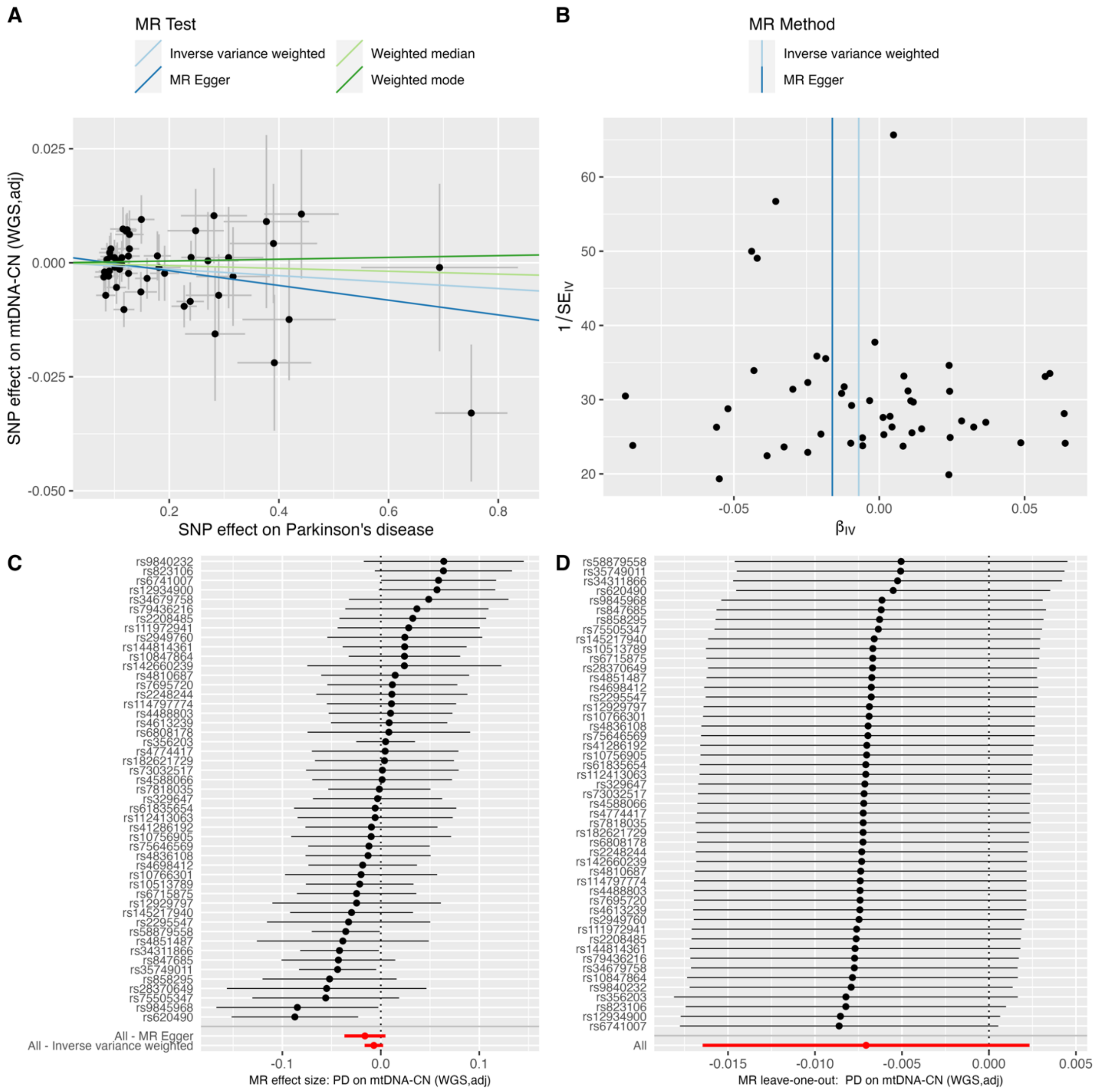


**Supplementary Fig. 16 Mendelian Randomization analysis of PD on mtDNA-CN (WGS, adjusted).**

**a**, scatter plots showing the causal association; **b**, funnel plots; **c**, single SNP effect size plot; **d**, leave-one-out plot.

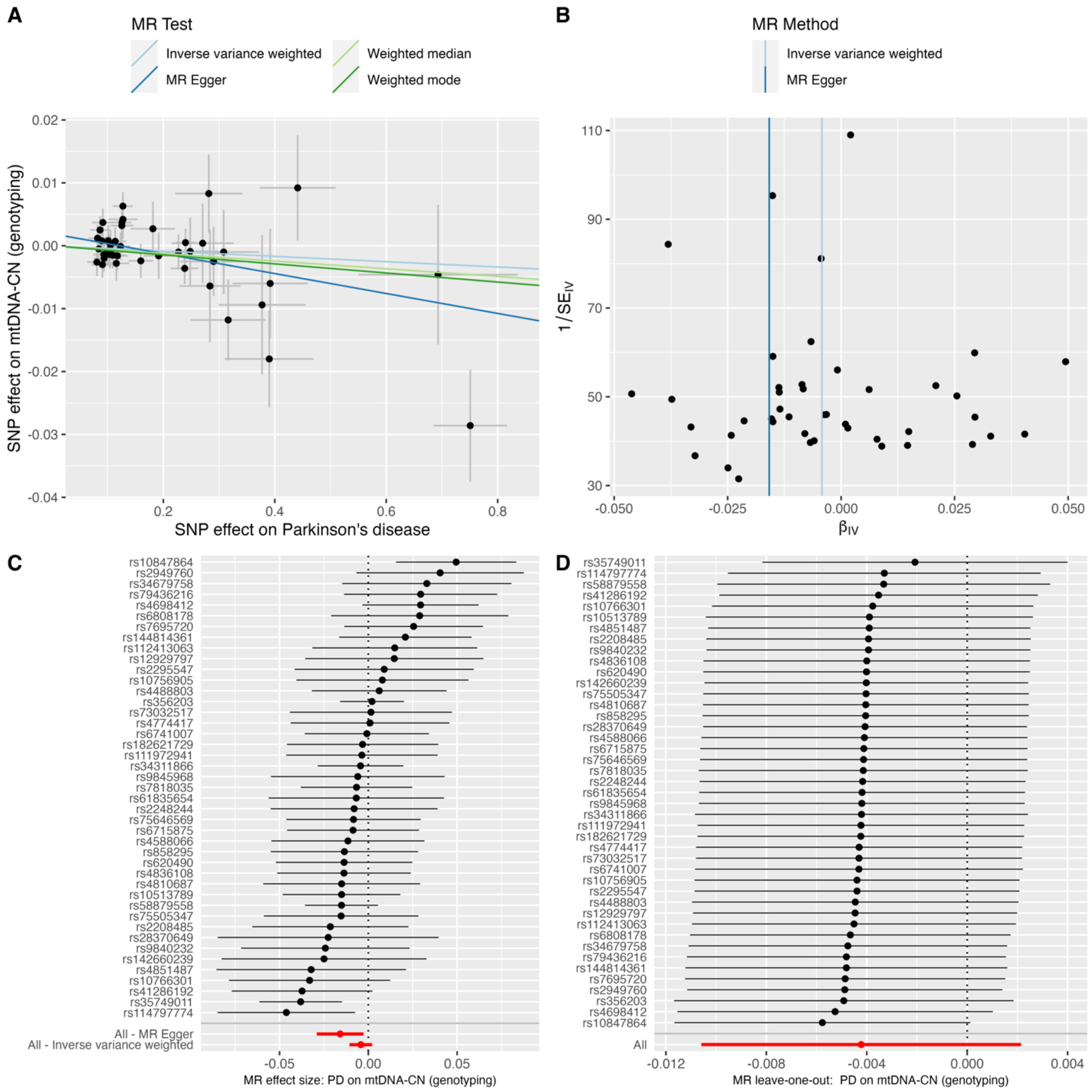


**Supplementary Fig. 17 Mendelian Randomization analysis of PD on mtDNA-CN (genotyping).**

**a**, scatter plots showing the causal association; **b**, funnel plots; **c**, single SNP effect size plot; **d**, leave-one-out plot.

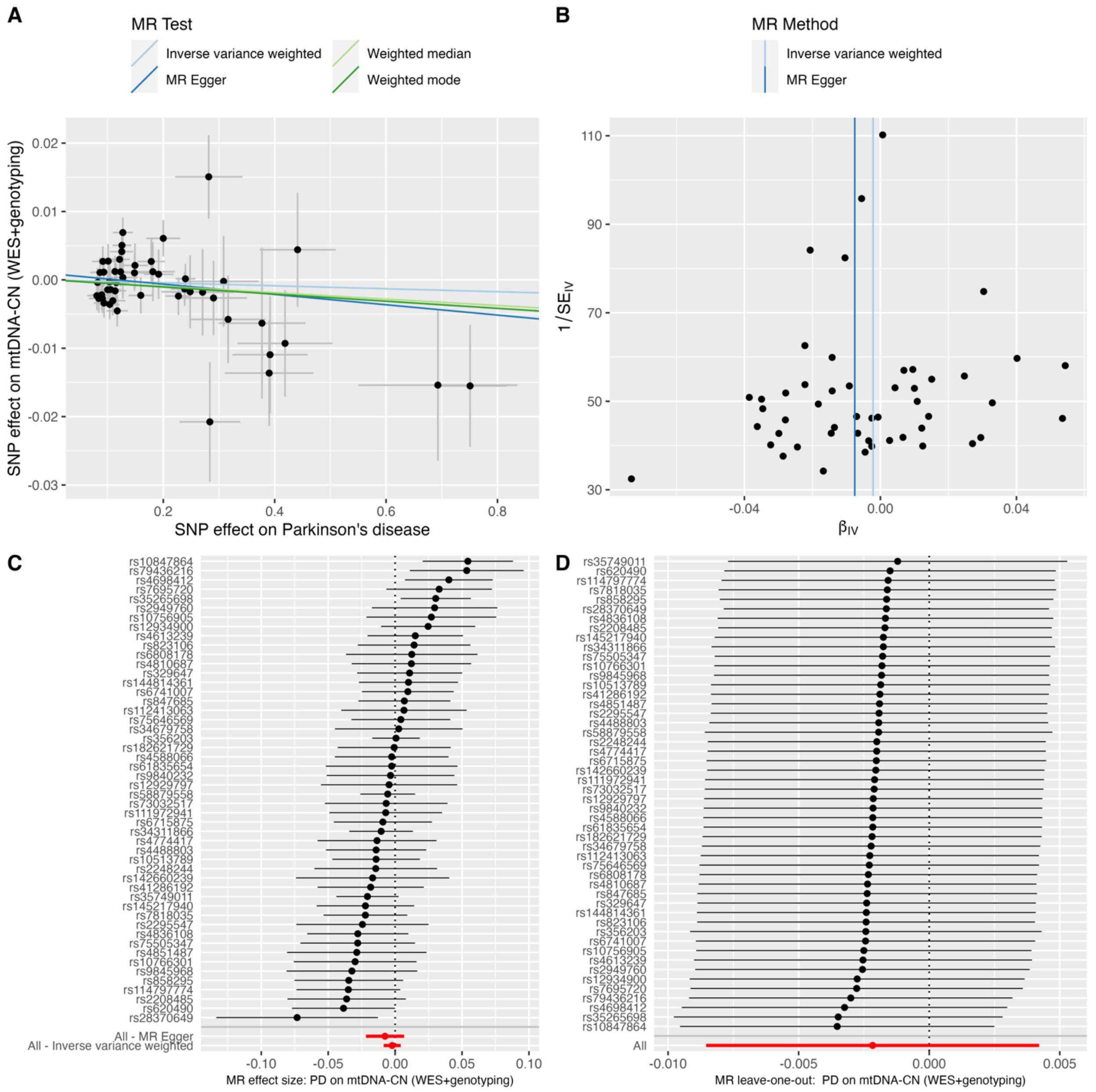


**Supplementary Fig. 18 Mendelian Randomization analysis of PD on mtDNA-CN (WES + genotyping).**

**a**, scatter plots showing the causal association; **b**, funnel plots; **c**, single SNP effect size plot; **d**, leave-one-out plot.

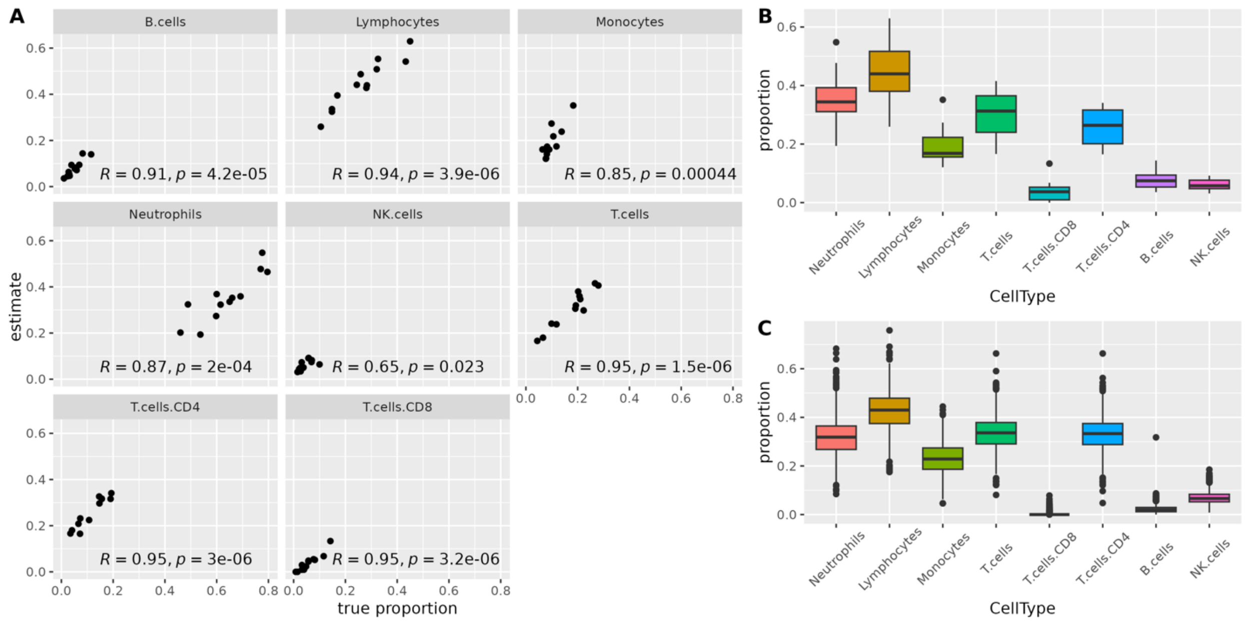


**Supplementary Fig. 19 Validation of CIBERSORTx cell type proportion estimates.**

**a**, Pearson correlation tests comparing estimated cell type proportions with ground truth proportions; **b**, boxplot displaying the range of estimated proportions for the validation cohort, consisting of whole blood bulk RNA-seq data from 12 healthy adults, in each of the eight major cell types; **c**, boxplot illustrating the range of estimated proportions for the bulk RNA-seq data from healthy controls in the AMP PD data. Lymphocyte proportions were derived with the cumulative proportions of T cells, B cells, natural killer (NK) cells.

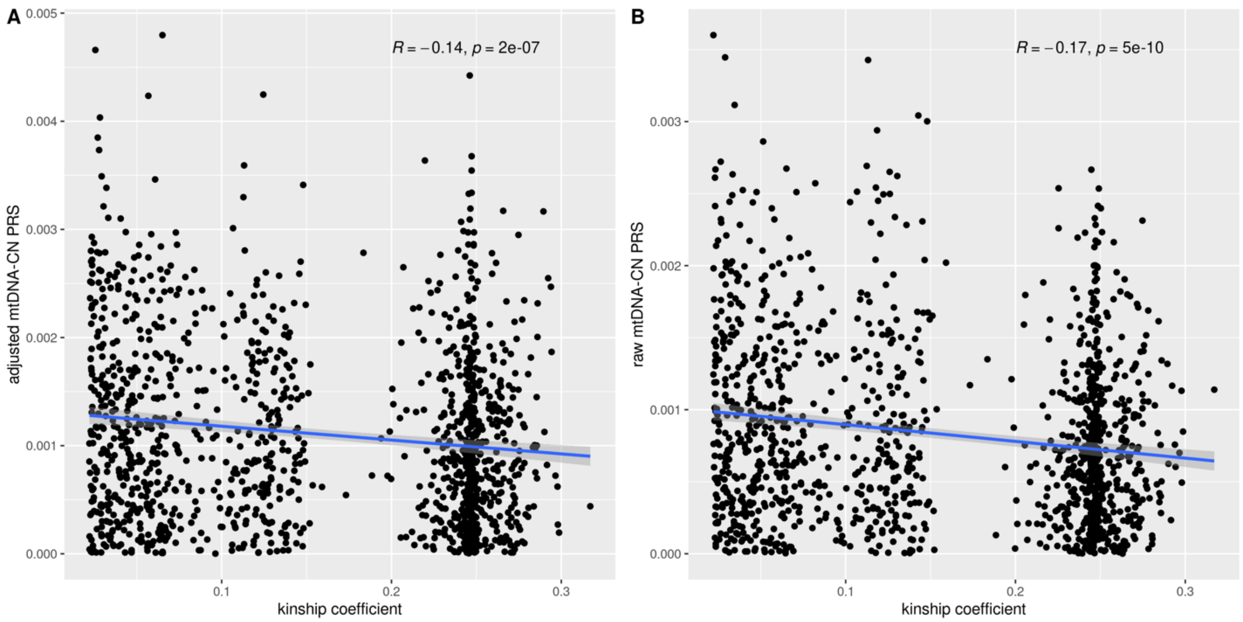


**Supplementary Fig. 20 Correlation analysis between mtDNA-CN PRSs and kinship coefficient.**

**a**, scatter plot shows the negative correlation between kinship coefficient and adjusted mtDNA-CN PRS (R = -0.14, *p* = 2e-07); **b**, scatter plot shows the negative correlation between kinship coefficient and raw mtDNA-CN PRS (R = -0.17, *p* = 5e-10).
